# Clinically feasible liver tumour cell size measurement through histology-informed *in vivo* diffusion MRI

**DOI:** 10.1101/2024.04.26.24306429

**Authors:** Francesco Grussu, Kinga Bernatowicz, Marco Palombo, Irene Casanova-Salas, Daniel Navarro-Garcia, Ignasi Barba, Sara Simonetti, Garazi Serna, Athanasios Grigoriou, Carlos Macarro, Anna Voronova, Valezka Garay, Juan Francisco Corral, Marta Vidorreta, Pablo García-Polo García, Xavier Merino, Richard Mast, Núria Rosón, Manuel Escobar, Maria Vieito, Rodrigo Toledo, Paolo Nuciforo, Joaquin Mateo, Elena Garralda, Raquel Perez-Lopez

## Abstract

Innovative diffusion Magnetic Resonance Imaging (dMRI) models enable the non-invasive measurement of cancer biological properties *in vivo*. However, while cancers frequently spread to the liver, models tailored for liver application and easy to deploy in the clinic are still sought. We fill this gap by delivering a practical, clinically-viable dMRI framework for liver tumour imaging, informing its design through histology. By comparing dMRI and histological data from mice and cancer patients, we select a dMRI signal model of restricted intra-cellular diffusion with negligible extra-cellular contributions, maximising radiological-histological correlations. The model enables non-invasive liver cancer phenotyping, providing cell size and density estimates that i) correlate with their histopathology counterparts, ii) are associated to cell proliferation and tumour volume, and iii) that distinguish tumour types. By delivering metrics that are biologically meaningful, our approach may complement standard-of-care radiology, and become a new tool for enhanced cancer characterisation in precision oncology.

## Introduction

The clinical use of Magnetic Resonance Imaging (MRI) in cancer is based on revealing the presence of tumours within an area of interest, and on measuring their size. Nonetheless, MRI also offers the possibility of obtaining estimates of biological properties within each pixel making up the tumour image. This approach, known as quantitative MRI (qMRI)^1^, involves the acquisition of multiple MRI contrasts, which are then analysed jointly with a mathetmatical model. qMRI provides promising metrics, which could become quantitative biomarkers complementing the qualitative assessment by the expert radiologist^2^. Among existing qMRI approaches, diffusion MRI (dMRI) sensitises the MRI signal to water diffusion in biological tissues^1,3,4^. Since the patterns of diffusion are influenced by the microenvironment where diffusion takes place (e.g., by obstacles such as cell membranes), dMRI ultimately enables the per-voxel estimation of properties related to the size and density of cells^5–7^, bridging the gap between radiology and histology.

dMRI has shown promise in multiple areas, including brain^5^, spinal cord^8^, prostate^6^, breast^9^, or liver imaging^10,11^. The liver is a frequent site for cancer metastasisation^12^, and liver tumours are common targets for treatment response assessment in oncology. However, current image-based response criteria such as RECIST^13^ have limitations, in that they rely on MRI or computed tomography merely to measure tumour size, without accounting for changes under therapy at the cellular level. Novel dMRI metrics could instead enable the non-invasive characterisation of cancer microenvironments, shedding light on the composition of tumours that cannot be biopsied. The new readouts could also provide information on tumour heterogeneity, relevant to fight treatment resistance^14,15^, and better stratify patients for personalised treatment planning, reducing sample sizes in clinical trials and ultimately improving patient outcomes^16^.

In practice, the estimation of the biological properties through dMRI is made possible by mathematical models, which are used to link the observed image contrasts to the biological properties themselves^17–20^. However, despite their promise, advanced liver dMRI models are relegated almost exclusively to research contexts, and seldom used in real-world clinical settings. This is due to two main reasons: i) to the high number of dMRI images (and hence long scan time) required to support the fitting of such models, and ii) to the requirement for specialised dMRI acquisitions^21^, beyond those made available by manufacturers in the scanner console.

In this study we aim to fill this clinical-research gap by delivering a practical liver dMRI framework that is truly feasible in hospital settings, i.e., on 1.5T or 3T systems, with scan time that does not exceed 15 minutes, and using vendor-provided dMRI acquisitions. To this end, we compared a set of candidate, state-of-the-art approachs using a rich data set of dMRI scans and hematoxylin and eosin (HE)-stained images, from excised mouse livers and patients’ liver biopsies. We used these unique data to select the approach maximising the correlation of radiological and histological estimates of metrics such as cell size, corroborating results with computer simulations. The proposed approach is based on a model of restricted, intra-cellular (IC) diffusion with negligible extra-cellular (EC) signal contributions, which is fitted to highly diffusion-weighted (DW) images. The model provides histologically-meaningful estimates of cell size and density, which are also shown to be associated to cell proliferation within liver tumours, as well as to the tumour size. These results suggest that our dMRI framework may provide novel, histologically-relevant descriptors of tumour characteristics. These may complement standard-of-care readouts in research and clinical practice, equipping oncologists with new tools for precision oncology in hospital settings.

## Results

### Overview: mouse and human data for histology-informed dMRI signal modelling

We designed and demonstrated our dMRI approach using data obtained in fixed mouse livers (*mouse data*) and in cancer patients *in vivo* (*human data*) (Fig. 1). Mouse data consists of pulsed gradient spin echo (PGSE) DW MRI scans of seven fixed mouse livers, performed *ex vivo* on a 9.4T Bruker system. It also includes whole-organ HE-stained sections, obtained at known radiographic position. We obtained livers of mice sacrificed as part of ongoing xenograft model development studies in prostate cancer. Six had been implanted with biopsies of prostate cancer patients, while one had not had any implantation. While the livers from the implanted mice did not grow any tumours, they feature a variety of pathologies, with three unique histopathological phenotypes (Fig. S1). The liver from the mouse with no implantation features normal liver structures, and we will refer to it as *Control*. Of the six implanted cases, two also show normal liver tissue, with normal representation of all hepatic structures. We will refer to these cases as *Pat_NA1_* and *Pat_NA2_* (patient biopsy implantation, but normal appearing). Another case exhibits generalised necrosis and diffuse acute and chronic inflammation surrounding necrotic areas, with presence of occluded thrombotic vessels. This specimen will be identified as *Pat_nec_* (patient biopsy implantation, with necrosis). Finally, three specimens feature an immature, lymphoproliferative process, with various degrees of infiltration of small, lymphoid, atypical cells with abundant mitosis, which infiltrate portal vessels and sinusoidal capillaries, but without producing tumours. These will be referred to as *Pat_inf1_* to *Pat_inf3_* (patient biopsy implantation, with lymphoid cell infiltration).

**Fig. 1.**
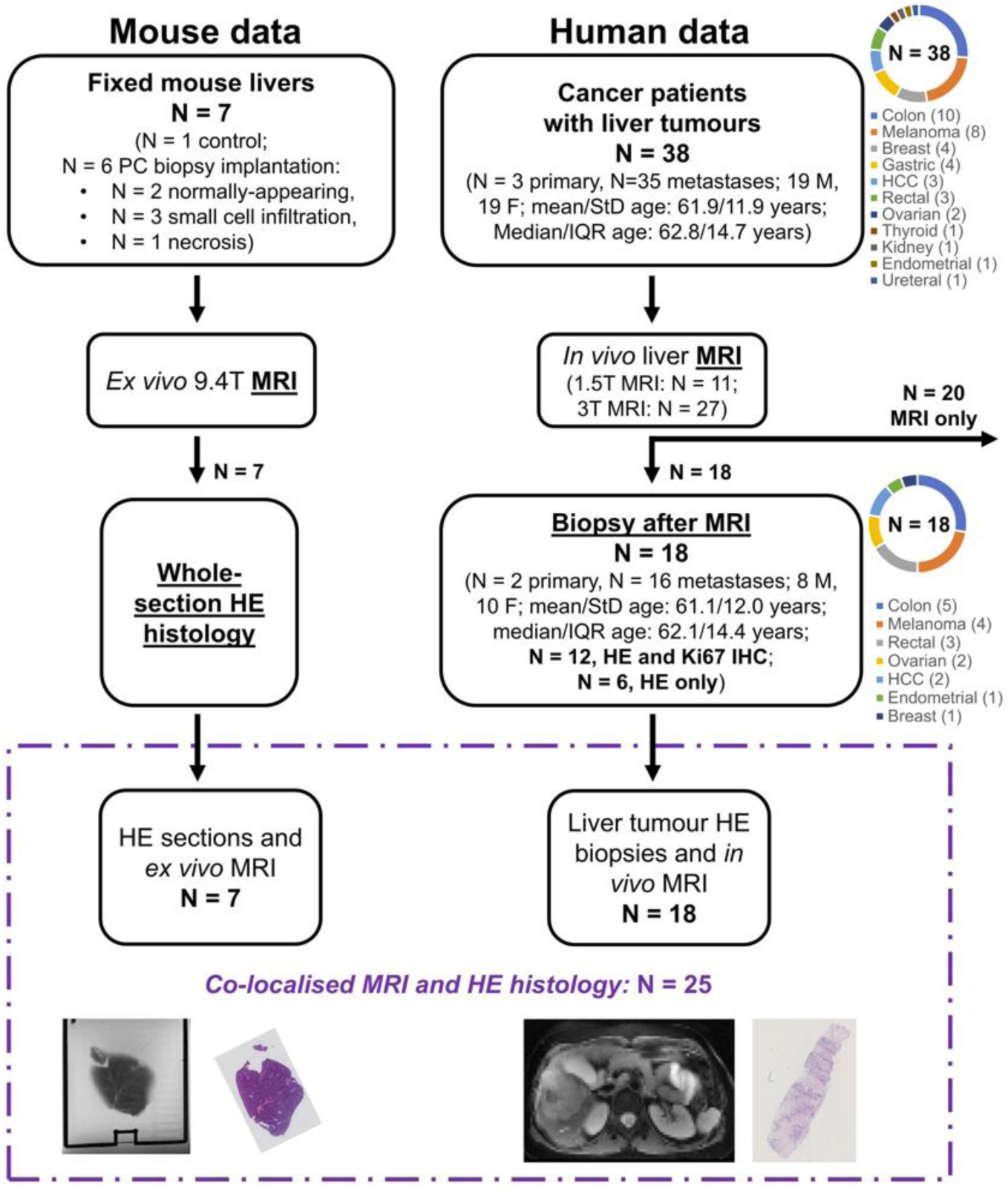
Illustration of the liver MRI and histology data used in the study. Our data set consisted of preclinical mouse data and clinical human data. The mouse data encompasses dMRI scans of seven fixed livers from mice (six implanted with tissue from biopsies of patients suffering from prostate cancer (PC); one without any implantation). We scanned the livers *ex vivo* on a 9.4T system, and obtained HE histological sections at known position. The clinical data includes *in vivo* liver dMRI scans performed on 38 patients suffering from advanced liver tumours. Scans were performed on clinical 1.5T and 3T MRI systems, and a HE-stained biopsy from one of the images tumours was collected after MRI in 18 cases. In 12 out of these 18, Ki67 immunohistochemsitry (IHC), demonstrating cell proliferation, was available beyond routine HE. cases In the figure, PC stands for prostate cancer; HCC for hepatocellular carcinoma; IHC for immunohistochemsitry.

We obtained human data on cancer patients suffering from advanced solid tumours, participating in an imaging study at the Barcelona Vall d’Hebron Institute of Oncology (VHIO, Spain), before being enrolled in a number of clinical trials. We included data from 38 patients with liver malignancies (mean/standard deviation (SD) of age: 61.88/11.90 years; median/inter-quartile range (IQR) of age: 62.8/14.7 years; 19 male, 19 female), of which 3 suffered from primary hepatocellular carcinoma (HCC), while 35 had liver metastases from different primary cancers (10 colon, 8 melanoma, 4 breast, 4 gastric, 3 rectal, 2 ovarian, 1 renal, 1 endometrial, 1 ureteral, 1 thyroidal). dMRI was based on diffusion-weighted (DW) echo planar imaging (EPI). We also obtained digitised HE-stained biopsies from one of imaged liver tumours approximately one week after MRI in 18 cases (mean/SD of age: 61.1/12.0 years; median/IQR of age: 62.1/14.4 years; 8 male, 10 female). Of these 18 patients, 2 suffered from primary HCC, while 16 had liver metastases (5 from colon cancer, 4 from melanoma, 3 from rectal cancer, 2 from ovarian cancer, 1 from breast and endometrial cancer). In 12 cases, Ki67 imunnohistochemsitry (IHC), demonstrating cell proliferation, was also available beyond routine HE stains.

### Overview: dMRI signal models for cell size and density measurement

We studied five biophysical dMRI models, grouped into two families (Fig. 2A). All models describe the dMRI signal as originating from a combination of IC and EC contributions^6,17,20^, making different assumptions. The first family, which is more general, includes:

i. *Diff-in-exTD*: a model accounting for restricted IC diffusion within spherical cells^6^, and hindered diffusion in the EC space, with diffusion time dependence^22^ in both IC and EC signals^23^. The diffusion time quantifies the time during which water molecules can sense cellular barriers, before the MR signal is measured.
ii. *Diff-in-ex*: similar to model *Diff-in-exTD*, but neglecting diffusion time dependence in the EC space. Popular body dMRI techniques such as IMPULSED^17^ or VERDICT^18^ are essentially implementations of this model.

**Fig. 2.**
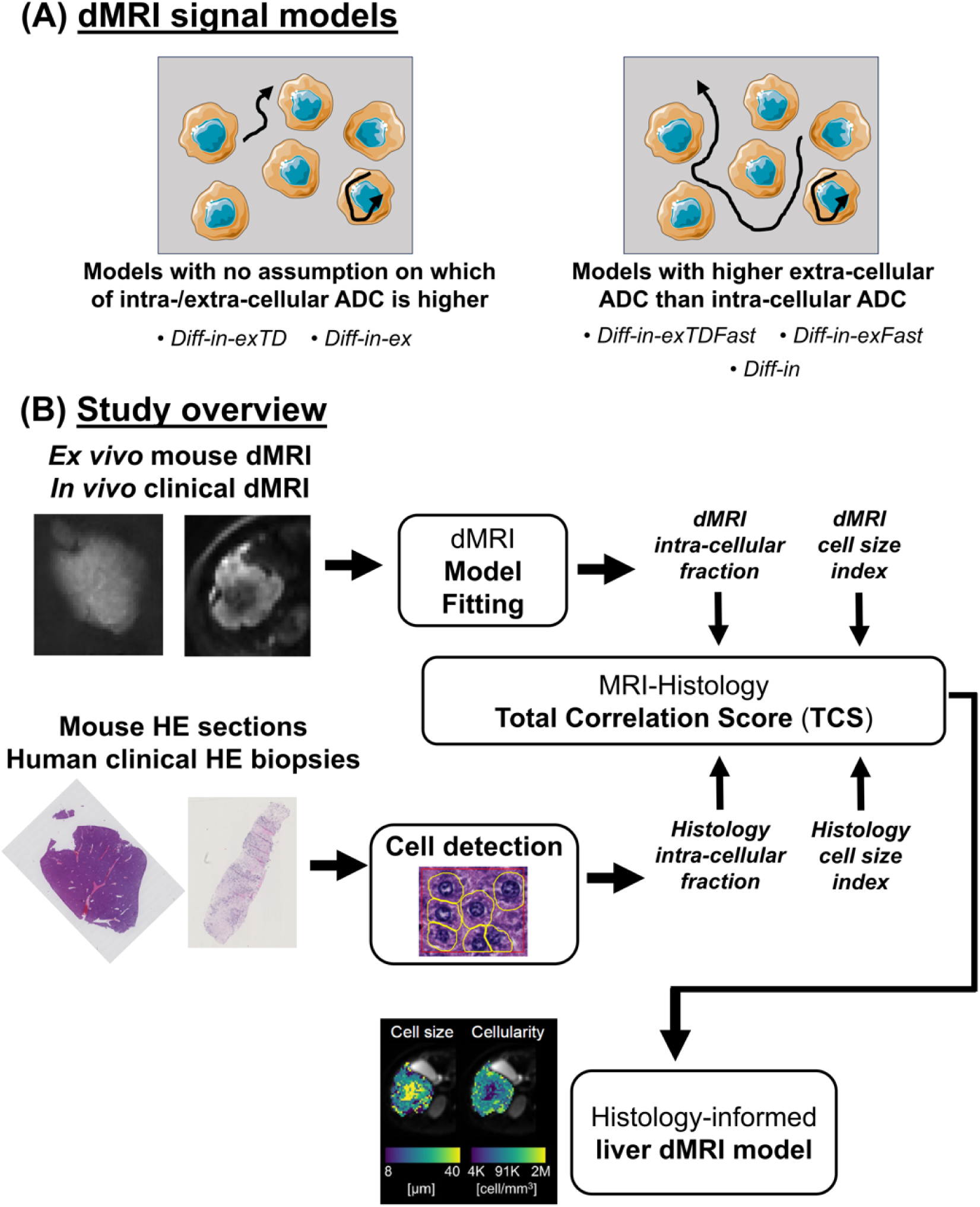
Description of the dMRI signal models and study overview. (**A**), top: cartoon illustrating the two families of dMRI models considered in this study, consisting of 1) models with no assumption of which of intra/extra-cellular ADC is higher, and 2) models where the extra-cellular ADC is hypothesised to be higher than the intra-cellular ADC. (**B**), bottom: study overview. We analysed dMRI data from fixed mouse livers and from cancer patients imaged *in vivo* to derive estimates of intra-cellular fraction and of cell size. In parallel, we processed histological material from the same tissues (whole-liver sections for the preclinical mouse data; biopsies from one of the imaged tumours for the clinical data), and derived the histological counterparts of such dMRI metrics. We compared dMRI and histological cell size and intra-cellular fraction to select the dMRI model featuring the best fidelity to histology. In Fig. 2, pictures from Servier Medical Art have been used. Servier Medical Art by Servier is licensed under a Creative Commons Attribution 3.0 Unported License (https://creativecommons.org/licenses/by/3.0).

Conversely, the second family explicitly assumes that the EC apparent diffusion coefficient (ADC) is higher in the EC than in the IC space, i.e., that *ADC*_*E*_ > *ADC*_*I*_, and assumption used in certain body dMRI techniques^24^. This family includes:

i. *Diff-in-exTDFast*: equivalent to *Diff-in-exTD*, but ensuring that *ADC*_*E*_ > *ADC*_*I*_.
ii. *Diff-in-exFast*: equivalent to *Diff-in-ex*, but ensuring that *ADC*_*E*_ > *ADC*_*I*_.
iii. *Diff-in*: a model where it is hypothesised that the EC extra-cellular signal is negligible with respect to the IC one due to its much larger ADC.

All models enable the estimation of *volume-weighted mean cell size* (*VCS*_*MRI*_, expressed in µm) and *intra-cellular signal fraction* (*F*_*MRI*_, dimensionless). These can be combined into an apparent *cell density per unit volume* (*CD*_*MRI*_ = *F*_*MRI*_/*VCS*^3^, expressed in cell/mm^3^)^18^. All models were fitted twice: to the complete set of images, or to highly diffusion-weighted (DW) images only (i.e., to high *b*-values), to minimse signal contributions from the EC space. For comparison, we also considered routine ADC (in µm^2^/ms) and apparent diffusion excess kurtosis *K* (dimensionless) from diffusion kurtosis imaging (DKI)^25^, since these are popular dMRI indices sensitive to cancer cellularity and easy to compute from short acquisitions^18,26^.

We processed HE-stained histological data with automatic cell detection^27^ to derive histological counterparts of the dMRI metric. The two were compared to inform the design of a dMRI signal model maximising radiological-histological correlations (Fig. 2B). The histological metrics were: *histological volume-weighted mean cell size* (*VCS*_*histo*_, in µm), *histological intra-cellular area fraction* (*F*_*histo*_, dimensionless), and *histological cell density per unit area* (*CD*_*histo*_, in cell/mm^2^). In patients for which Ki-67 ICH was available, the fraction of histological material stained for Ki-67 within the biopsy was also computed through custom routines (metric *F*_*Ki*67_).

### A model of restricted intra-cellular diffusion with negligible extra-cellular contributions maximises radiological-histological correlations

We compared dMRI models through an MRI-Histology *Total Correlation Score* (TCS). TCS measures the overall correlation between histological and radiological readouts of cell size and intra-cellular fraction, and is obtained by summing Pearson’s correlation coefficients between MRI and histological cell size and IC fraction (i.e., *VCS*_*MRI*_and *VCS*_*histo*_, and *F*_*MRI*_and *F*_*histo*_). Negative correlations are penalised, since the model providing the highest TCS should be preferred. TCS values unequivocally suggest that models where the IC ADC is constrained to be smaller than the EC ADC provide higher correlations with histology. Among all dMRI implementations, fitting a model of IC restricted diffusion with negigilble EC signal contributions to highly DW images (referred to as model *Diff-in*) provides the highest TCS values, and hence the highest correlation to histological readouts (Fig. 3). Fig. 3A summarises the different dMRI models, while Fig. 3B and Fig. 3C report actual TCS values. In the figure, models where *ADC*_*E*_ > *ADC*_*I*_, shown in violet shades, provide consistently higher TCS values than models that do not make such an assumption (orange shades), with the highest TCS observed for model *Diff-in* fitted to highly DW images.

**Figure 3.**
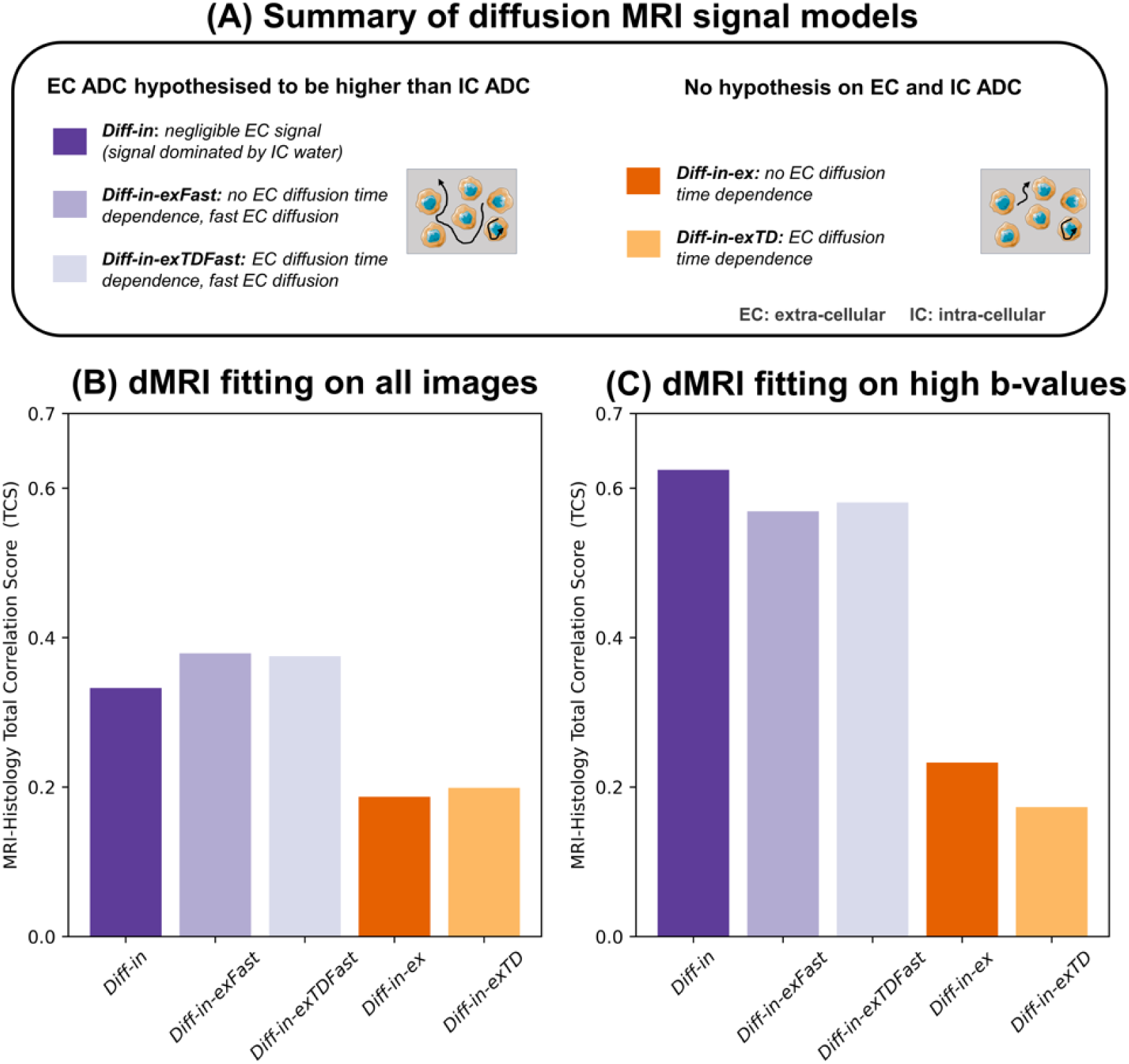
Biophysical dMRI signal model selection based on the MRI-histology Total Correlation Score (TCS). **(A)**: panel summarising the salient differences between the biophysical dMRI models compared in this study. Models can be divided in two families, i.e.: i) models where it is hypothesised that the extra-cellular ADC is higher than the intra-cellular ADC, and ii) models with no hypothesis on which, between intra-/extra-cellular ADC is higher. Violet shades are used for the first family (models *Diff-in*, *Diff-in-exFast* and *Diff-in-exTDFast*), while orange shades for the second family (models *Diff-in-ex* and *Diff-in-exTD*). **(B)**: values of TCS for all models, as obtained by fitting them on the whole set of images featuring negligible vascular signal contribution (*b* > 1000 s/mm^2^ in the fixed mouse livers; *b* > 100 s/mm^2^ *in vivo*). **(C)**: TCS values for all models when these are fitted only on high b-value images (*b* > 1800 s/mm^2^ in the fixed mouse livers; *b* > 900 s/mm^2^ *in vivo*). In this figure, pictures from Servier Medical Art have been used. Servier Medical Art by Servier is licensed under a Creative Commons Attribution 3.0 Unported License (https://creativecommons.org/licenses/by/3.0).

Additional model selection criteria used to compare dMRI models confirm findings from TCS-based model selection. The additional criteria were: an *Histology Fidelity Criterion* (HFC), measuring the sum of absolute errors in *F* and *VCS* estimation via dMRI, compared to histology; and the *Bayesian Information Criterion* (BIC)^28^. BIC is a common criterion for dMRI model selection^29,30^: it quantifies the model fitting quality, penalising model complexity. Note that model performance increases as both HFC and BIC decrease. Fig. S2 reports the number of times, in percentage terms, that a model provides the lowest HFC and BIC across our N = 25 MRI-histology cases. TCS ranking is confiremd: models hypothesising *ADC*_*E*_ > *ADC*_*I*_ are selected more frequently than models that do not do, according to HFC. The model *Diff-in* is the most selected model according to both BIC and HFC (Fig. S3.B; fitting to high b-value images). Fig. S3 splits HFC and BIC rankings depending on the MRI scanner. In all cases, models with *ADC*_*E*_ > *ADC*_*I*_ (*Diff-in*, *Diff-in-exFast*, *Diff-in-exFastTD*) are selected more frequently than models *Diff-in-ex* and *Diff-in-ex-TD*. When fitting is performed only on high b-value images, *Diff-in* is the most selected model according to both BIC and HFC.

### Computer simulations confirm results from MRI-histology model selection

We validated the model selection performed on mouse and human dMRI data thourgh computer simulations. These consisted in: i) synthesing dMRI signals for the three acquisition protocols used in this study, through Monte Carlo simulations; ii) fitting all candidate models to the synthetic dMRI signals; iii) selecting the best approach according to the TCS, HFC and BIC criteria introduced above. In our synthetic data, all model selection criteria confirm the selection of model *Diff-in* fitted to highly DW images, previously obtained on actual MRI-histology data. Fig. S4 shows the virtual tissues used to generate synthetic dMRI signals, i.e., made of packed spheres representing cells, as common in body dMRI^6,17,20,21^. Supplementary Tables S1, S2, and S3 report synthetic TCS, HFC and BIC rankings for all dMRI protocols. In the vast majority of cases, model *Diff-in* fitted to highly DW images yields the best estimation of cell size and IC fraction, thus corroborating results from *ex vivo* and *in vivo* dMRI.

### Cell size and density estimates from the proposed dMRI model correlate with histology

In view of the results obtained from the analysis of *in silico*, *ex vivo* and *in vivo* MRI data, our recommended liver tumour dMRI approach is the *fitting of model “Diff-in” – a one-compartment model of restricted, IC diffusion within spherical cells, with negligible EC signal contributions – to high b-values images* (≳ 1800 s/mm^2^ *ex vivo*, ≳ 900 s/mm^2^ *in vivo*).

The model provides estimates of IC fraction, cell size and cell density that are weakly, moderately and strongly correlated to the underlying histology. Table 1 shows Pearson’s correlation between *Diff-in* metrics *F*_*MRI*_ (intra-cellular fraction), *VCS*_*MRI*_(cell size index) and *CD*_*MRI*_(cell density) with their histology counterparts *F*_*histo*_, *VCS*_*histo*_and *CD*_*histo*_. We observe a weak correlation between *F*_*MRI*_ and *F*_*histo*_ (*r* = 0.19, *p* = 0.370), moderate between *VCS*_*MRI*_ and *VCS*_*histo*_ (*r* = 0.44, *p* = 0.029), and strong between *CD*_*MRI*_and *CD*_*histo*_(*r* = 0.70, *p* = 0.0001). The weak correlation between *F*_*MRI*_and *F*_*histo*_is likely due, at least in part, to the fact that *F*_*MRI*_is a signal fraction, rather than an actual volume/area fraction. This implies that *F*_*MRI*_ also encodes T2/T1 differences between IC and EC signals, unlike *F*_*histo*_^31^. Moreover, *F*_*MRI*_values may have also been biased by the exchange between intra- and extra-cellular water^19,32^, especially in presence of small cells^19^ – a phenomenon that is is not accounted for in none of the studied signal models. Finally, we emphasise that in the N = 18 human observations, dMRI metrics from an entire tumour have been compared to a small sliver of biopsied tissue. This implies that the correlation between dMRI and histology may have been underestimated, especially for the IC fraction, for example when the biopsy misses large areas of necrosis or fibrosis, which affect *F*_*MRI*_ values.

**Table 1.** Correlation between metrics from the selected biophysical dMRI approach with their direct histological counterparts. The table reports Pearson’s correlation coefficients *r* and corresponding p-values *p* for dMRI metrics *F*_*MRI*_ (intra-cellular fraction), *VCS*_*MRI*_ (volume-weighted cell size index) and *CD*_*MRI*_(cell density per unit volume) of model *Diff-in*, fitted to high b-value images. The correlations are reported between each MRI metric and its direct histological counterpart (*F*_*histo*_for *F*_*MRI*_; *VCS*_*histo*_for *VCS*_*MRI*_; and *CD*_*histo*_for *CD*_*MRI*_). For comparison, the table also reports correlation coefficients between routine ADC and *K* from DKI and each of *F*_*histo*_, *VCS*_*histo*_and *CD*_*histo*_. The sample size was N = 25, implying that *p* < 0.05 if | *r* | > 0.396. *p* < 0.05 is flagged by bold font.

| | Histology $F_{histo}$ | Histology $vCS_{histo}$ | Histology $CD_{histo}$ |
| --- | --- | --- | --- |
| **Model *Diff-in* fitted to high $b$-value images** | With $F_{MRI}$<br>$r = 0.19$; $p = 0.370$ | With $vCS_{MRI}$<br>**$r = 0.44$; $p = 0.029$** | With $CD_{MRI}$<br>**$r = 0.70$; $p = 0.0001$** |
| **DKI $ADC$** | $r = -0.28$; $p = 0.180$ | **$r = 0.49$; $p = 0.014$** | **$r = -0.47$; $p = 0.017$** |
| **DKI $K$** | **$r = 0.40$; $p = 0.048$** | $r = -0.31$; $p = 0.130$ | **$r = 0.43$; $p = 0.033$** |

Table 1 also reports correlation coefficients for DKI ADC and kurtosis *K*. The correlations are in line with previous studies^33,34^. For example, both ADC and *K* exhibit significant, moderate correlations with histological properties, i.e., a negative correlation with cell density *CD*_*histo*_for ADC, and a positive correlation with *CD*_*histo*_for *K* (*r* = –0.47 and 0.43 respectively). Significant correlations are also seen between DKI metrics and *F*_*histo*_ (*r* = 0.40, *p* = 0.048 for *K*).

Fig. S5 and Fig. S6 show Pearson’s correlation coefficients for all possible pairs of metrics, in the form of correlation matrices. Supplementary Data S1 and S2 contain all MRI/histology metric values used to generate such matrices. In general, metrics from dMRI models where *ADC*_*E*_ > *ADC*_*I*_ show stronger correlations with their histological counterparts than models *Diff-in-exTD* and *Diff-in-ex*. Specifically, we observe the strongest dMRI-histology correlations for model *Diff-in* fitted to high b-value images. Correlations among pairs of dMRI metrics are also seen, e.g., a strong negative correlation between *CD*_*MRI*_and *VCS*_*MRI*_(*r* = –0.81 for model *Diff-in* fitted to high b-value images). This finding, indicating that tighter cell packings per unit volume are achieved with smaller cells, is mirrored by histological *CD*_*histo*_ and *VCS*_*histo*_ (*r* = –0.88 between these two), and thus appears biophysically plausible.

### The proposed dMRI model reveals histologically-meaningful tumour characteristics

The proposed model *Diff-in* is demonstrated in 7 fixed livers of mice suffering from a variety of pathologies, as well as in a cohort of 38 patients suffering from advanced solid tumours of the liver (both primary and metastatic). Voxel-wise *Diff-in* parameteric maps reveal intra-tissue and inter-tissue contrasts that are histologically meaningful, being compatible with histological features observed in HE stains. These include, for example, presence of small cells in areas characterised by lymphoproliferative processes in mouse livers, or reductions in cell density in necrosis or fibrosis in human tumours.

Fig. 4 shows *Diff-in* and histological maps in 3 mouse livers, representative of the 3 phenotypes seen in our mouse data (*Control*, for normal liver structures; *Pat_inf1_*, for small cell infiltration; *Pat_nec_*, for necrosis). Visually, we observe excellent co-localisation between MRI slices and HE sections. The histological details reveal higher cellularity in sample *Pat_inf1_* compared to *Control*, due to packing of small cells in between larger hepatocytes, or an alternation of areas with lower/higher cell density in sample *Pat_nec_*. These qualitative trends are confirmed in the histological maps *F*_*histo*_, *VCS*_*histo*_, *CD*_*histo*_, with values in physiologically plausible ranges, as for example intra-cellular fractions around 0.75 and cell sizes of the order of 20 µm^35,36^. Maps *F*_*MRI*_, *VCS*_*MRI*_and *CD*_*MRI*_replicate the contrasts seeing in their histological counterparts *F*_*histo*_, *VCS*_*histo*_ and *CD*_*histo*_. Fig. S7 shows standard DKI ADC and kurtosis excess *K* in the same mouse livers. Visual trends highlight that the higher cell density of sample *Pat_inf1_* translates to remarkably reduced ADC and increased *K* compared to the *Control*. Lastly, Fig. S8 and S9 show additional dMRI metrics, including metrics from model *Diff-in-exFast* (i.e., the one providing the higher TCS when fitting is performed to the whole dMR image set). The figures highlight that overall, spatial trends seen in maps from the selected model *Diff-in* agree with those seen in *Diff-in-exFast*, but metrics from the latter appear noisier. Metrics *D*_0,*I*_ and *D*_*E*,∞_ show limited between-sample contrast, and are difficult to validate histologically. Table S4 report qualitative per-sample mean and standard deviation of all MRI and histology metrics in mice. *F*_*MRI*_ slightly underestimates *F*_*histo*_, while *VCS*_*MRI*_slightly overestimates *VCS*_*histo*_. We speculate that the discrepancies may be due, at least in part, to unaccounted factors such as variability in intrinsic cell shape/cytosol diffusivity^37^ or water exchange^19^, and by the difficulty of relating accurately 2D histology to 3D MRI^38^.

**Fig. 4.**
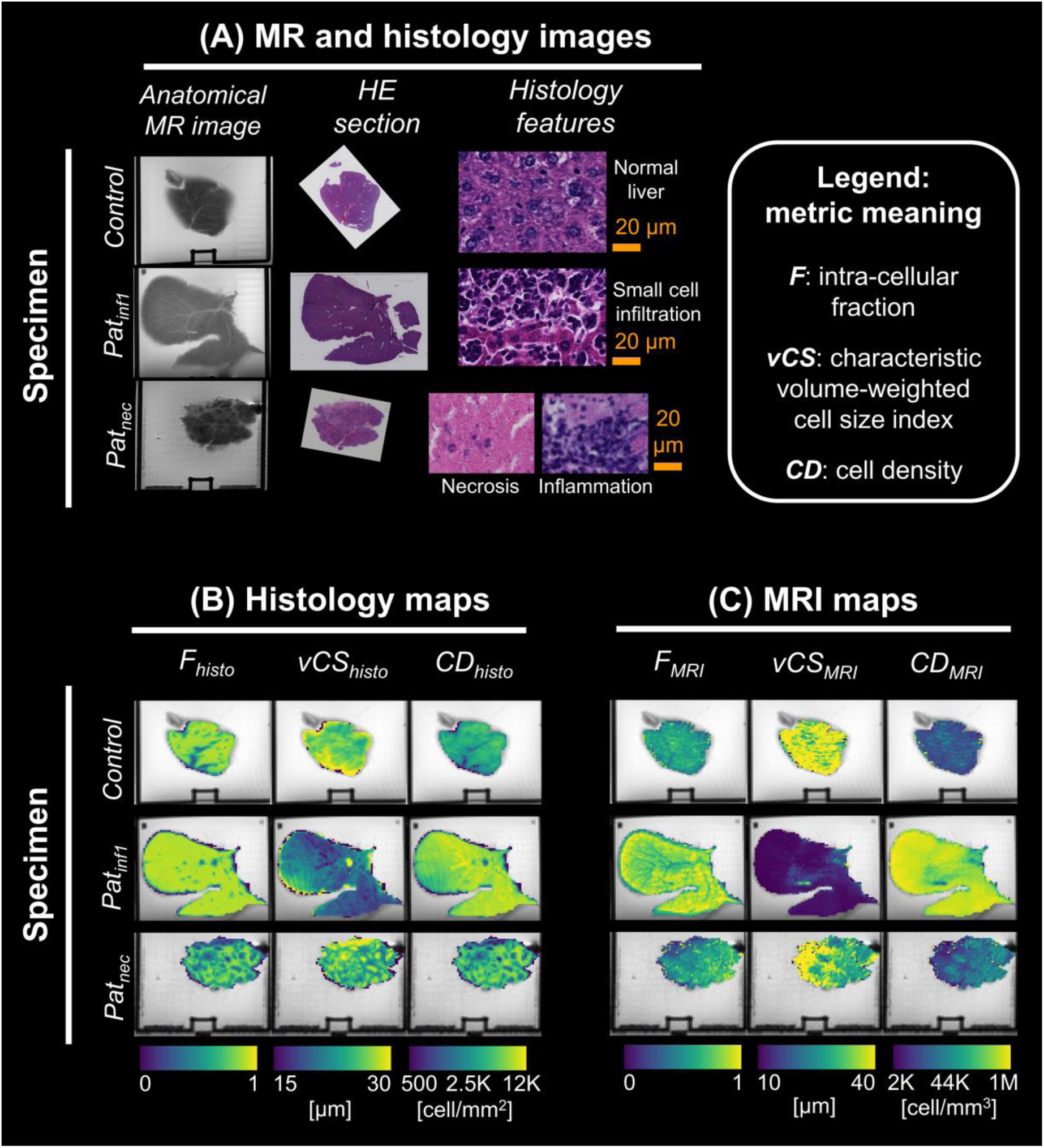
Maps from the selected dMRI model *Diff-in* with their histological counterparts in the fixed mouse livers scanned at 9.4T *ex vivo*. The figure reports MRI and histology data for 3 specimens, representative of the 3 microstructural phenotypes observed in our *ex vivo* data set, namely: normal liver structures (*Control* case); a proliferative process, characterized by infiltration of small cells (*Pat_inf1_* case); necrosis and inflammation (*Pat_nec_* case). For all specimens, the following is shown. **(A)**, *top left*: a high-resolution T2-w anatomical scan is shown next to the corresponding HE section, with histological details. **(B)**, *bottom left*: histological maps warped to the dMRI space (intra-cellular patch area fraction *F*_*histo*_; volume-weighted mean cell size index *VCS*_*histo*_; cell density per unit patch area *CD*_*histo*_). **(D)**, *bottom right*: dMRI maps *F*_*MRI*_, *VCS*_*MRI*_ and C*D*_*MRI*_ from the selected dMRI signal model (model *Diff-in*, fitted to high *b*-value images, i.e., *b* > 1800 s/mm^2^).

Fig. 5 shows *F*_*MRI*_, *VCS*_*MRI*_ and *CD*_*MRI*_ maps in patients, alongside biopsies. Histopathological assessment highlights the variety of characteristics that can coexist within advanced solid tumours, e.g.: areas of fibrosis; localised areas of tightly packed cancer cells, sourrounded by stromal fibres; necrosis. dMRI *F*_*MRI*_, *VCS*_*MRI*_and *CD*_*MRI*_show contrasts that are plausible with such histopathological features. For example, in a breast cancer liver metastasis in Fig. 5, we observe a core of low intra-cellular fraction *F*_*MRI*_ and low cell density *CD*_*MRI*_, compatible with necrosis. In a HCC case instead, we see areas of high *F*_*MRI*_ and high *CD*_*MRI*_, sourrounded by lower *F*_*MRI*_ and lower *CD*_*MRI*_, potentially indicating the alternation of high cell densities with fibrotic tissue. Fig. S10 shows routine dMRI *ADC* and *K* in the same tumours. These show plausible contrasts given the known tumour histology, e.g., a core of high *ADC* and low *K* are seen in the necrotic core of the breast cancer tumour. Supplementary Fig. S11 shows *F*_*MRI*_, *VCS*_*MRI*_ and *CD*_*MRI*_from model *Diff-in-exFast*. Image contrasts match visually those seen in *Diff-in* metrics (the proposed approach), although *Diff-in-exFast* metrics appear noisier on visual inspection. Fig. S12 shows intra-cellular cytosol diffusivity *D*_0,*I*_ asymptotic *ADC*_*E*_ (*D*_*E*,∞_). Their speckled appearance suggests that these metrics are difficult to measure accurately *in vivo*^17,39^.

**Fig. 5.**
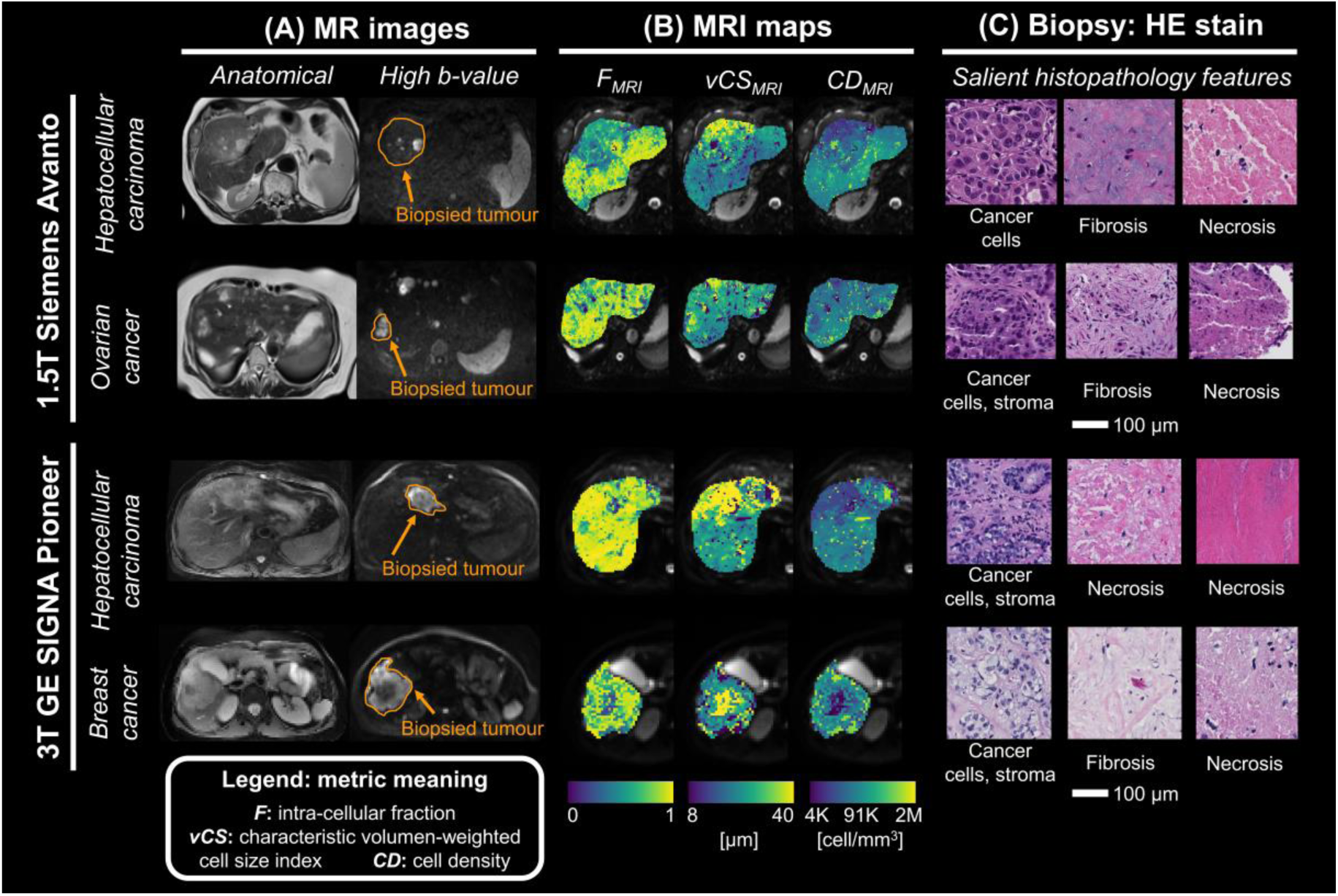
Examples of maps from the proposed dMRI model *Diff-in* in liver tumours of patients scanned at 1.5T and 3T *in vivo*, with co-localised biopsies. MRI maps are shown in a biopsied liver tumour in two patients for each MRI scanner, arranged along rows. **(A)**: examples of slices from the high-resolution anatomical T2-w image and from a high b-value image, with biopsied tumour outlined. **(C)**: maps from the selected model (*Diff-in*, fitted to high b-value images *b* > 900 s/mm^2^). From left to right: intra-cellular signal fraction F_MRI_; volume-weighted mean cell size index *VCS*_*MRI*_; cell density per unit volume C*D*_*MRI*_. **(C)**: histological details from the HE-stained biopsy. For the 1.5T Siemens scanner (first and second rows from top) we report: patient 6 (primary hepatocellular carcinoma) and patient 3 (liver metastases from ovarian cancer). For the 3T GE scanner (third and fourth rows from top) we report: patient 24 (primary hepatocellular carcinoma (HCC)) and patient 30 (liver metastases from breast cancer).

### Metrics from the proposed dMRI model correlate with cell proliferation in liver tumours

We tested the potential utility of the proposed model for non-invasive liver tumour phenotyping. To this end, we studied associations between dMRI metrics an cell proliferation within patients’ biopsied liver tumours, quantified through Ki67 immunohistochemistry (IHC). The maing finding of this analysis is that dMRI cell size (*VCS*_*MRI*_) and density (*CD*_*MRI*_) are respectively negatively and positively associated to the fraction of Ki67 staining (*F*_*Ki*67_). This results, confirmed by histological cell size and density (*VCS*_*histo*_and *CD*_*histo*_), suggests that tumours featuring high cell proliferation are made, on average, by smaller and more densely packed cells, compared to tumours with lower proliferation. Fig. 6 visualises the association between *F*_*Ki*67_ and MRI/histological cell size and density through scatter plots, reporting Pearons’s correlation coefficients. There is a strong, positive correlation between *CD*_*MRI*_ and *F*_*Ki*67_ (*r* = 0.80, *p* = 0.006), which is confirmed by a positive correlation between *CD*_*histo*_ and *F*_*Ki*67_(*r* = 0.66, *p* = 0.038). Regarding cell size, there is a moderate, negative correlation between *VCS*_*MRI*_and *F*_*Ki*67_(*r* = – 0.53, *p* = 0.111). This correlation, albeit not statistically significant owing to the small sample size (N = 10), is in the same direction of the correlation observed between histological *VCS*_*histo*_ and *F*_*Ki*67_ (*r* = –0.68, *p* = 0.031). Concerning other MRI metrics, weaker correlations are seen between DKI indices and *F*_*Ki*67_(*r* = 0.17, *p* = 0.633 between *ADC* and *F*_*Ki*67_; *r* = 012, *p* = 0.744 between *K* and *F*_*Ki*67_), or between *F*_*MRI*_ and *D*_0,*I*_ from the proposed dMRI model and *F*_*Ki*67_ (*r* = –0.35, *p* = 0.319 between *F*_*MRI*_ and *F*_*Ki*67_; *r* = 0.07, *p* = 0.857 between *D*_0,*I*_ and *F*_*Ki*67_).

**Fig. 6.**
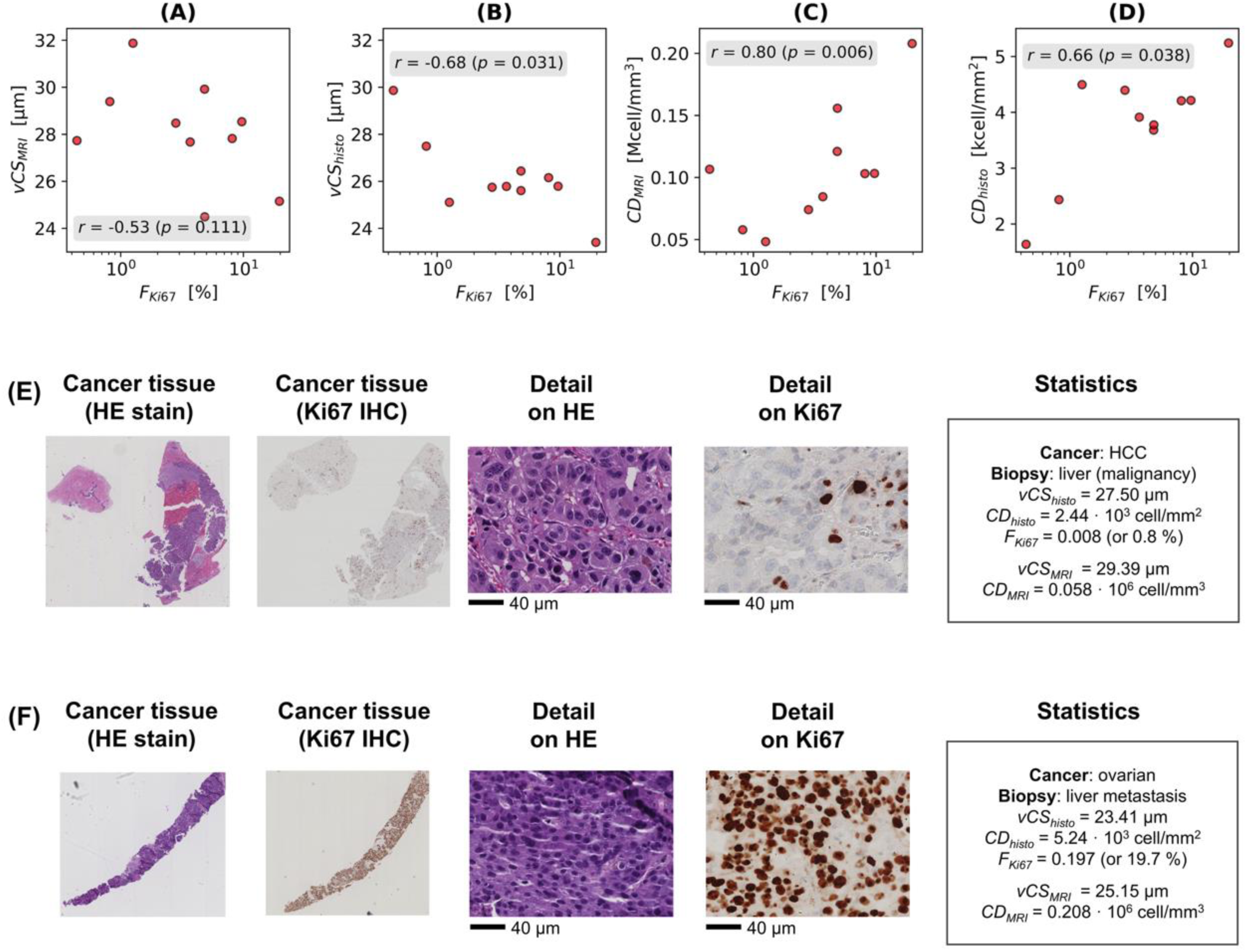
Association between dMRI metrics and cell proliferation within the biopsied tumours in patients. Top row: scatter plots and Pearson’s correlation coefficients between *F*_*Ki*67_ (strength of the IHC stain demonstrating proliferation within the biopsied tumour, here expressed in percentage terms) and indices of cell size and density from both MRI and HE histology (sample size: N = 10). From left to right, **(A):** scatter plot and correlation between *VCS*_*MRI*_ and *F*_*Ki*67_; **(B):** between *VCS*_*histo*_ and *F*_*Ki*67_; **(C):** between *CD*_*MRI*_and *F*_*Ki*67_; **(D):** between *CD*_*histo*_and *F*_*Ki*67_. **(E)**, middle row: example of HE and Ki67 stains from a slowly-proliferating tumour (primary hepatocellular carcinoma, *F*_*Ki*67_of 0.8%). **(F)**, bottom row: example of HE and Ki67 stains from a fast-proliferating tumour (ovarian cancer liver metastasis, *F*_*Ki*67_of 19.7%), with a summary of salient MRI and histological metrics. MRI maps of the two patients are shown in Fig. 5 and Fig. 7 (both patients were scanned on the 1.5T system).

Fig. 6 also visualises examples of biopsies from two liver tumours, featuring respectively strong and weak Ki67 staining for cell proliferation. These are: a metastatic ovarian cancer (Fig. 6F), whose *F*_*Ki*67_is of 0.197, and a primary HCC (Fig. 6E), which features a *F*_*Ki*67_of 0.008. The former tumour, with stronger Ki67 immunostain, is characterised by smaller and more densely packed cells than the latter, as suggested by both MRI and histology (for histology: *VCS*_*histo*_ of 23.41 µm against 27.50 µm, and *CD*_*histo*_of 5.24 ‧ 10^3^ cell/mm^2^ against 2.44 ‧ 10^3^ cell/mm^2^; for MRI: *VCS*_*MRI*_ of 25.15 µm against 29.39 µm, and *CD*_*MRI*_ of against 2.08 ‧ 10^5^ cell/mm^3^ against 0.58 ‧ 10^5^ cell/mm^3^).

### Metrics from the proposed dMRI model are associated with macroscopic tumour characteristics

We investigated whether the microscopic composition of each individual liver tumour assessed through dMRI can explain its macroscopic phenotype, e.g., salient characteristics such as its volume. For this analysis, we studied correlations between tumour-wise dMRI indices and the corresponding tumour volume, using data from N = 140 liver tumours from the 38 patients. We also performed cross-validation experiments in which we predicted liver tumour volume through statistical models that used dMRI metrics as predictors. The main finding of this analysis is that *Diff-in* metrics are significantly correlated with tumour volume, as is the case, for example, for MRI-derived cell size *VCS*_*MRI*_. Statistical models that rely on cell size *VCS*_*MRI*_ as a regressor enable meaningful predictions of tumour volumes, which are significantly correlated with ground truth values, and outperforming predictions from statistical models based on DKI metrics.

Fig. 7A reports Pearson’s correlation coefficients between tumour volume and dMRI metrics from both *Diff-in* and DKI dMRI. The strongest, statistically significant associations are seen for *Diff-in* metrics *D*_0,*I*_and *VCS*_*MRI*_(*r* = –0.305, *p* = 2.50 ‧ 10^-4^ and *r* = 0.256, *p* = 2.26 ‧ 10^-3^ respectively), while no significant associations are seen for DKI ADC (*r* = –0.092, *p* = 0.277) and kurtosis *K* (*r* = –0.035, *p* = 0.680). Fig. 7B shows an example of two tumours, one larger (a primary hepatocellular carcinoma, with a volume of circa 431 ‧ 10^3^ mm^3^) and one considerably smaller (an ovarian cancer liver metastasis, with a volume of approximately 21 ‧ 10^3^ mm^3^) (same tumours shown in Fig. 6). Histology shows that the former contains, on average, larger cells than the latter, a contrast that is detected by dMRI *VCS*_*MRI*_. Fig. 7C to Fig. 7F report scatter plots of different pairs of dMRI metrics coloured by tumour volume, visualising the latter as a bi-dimensional function of the diffusion metrics. Trends are seen in the colouring of the plots, e.g., in panel 7F, where the dependence of tumour volume on *D*_0,*I*_and *VCS*_*MRI*_is visualised (Fig. 7F). In Fig. 7F, the tumour volume does not appear to be monotonic in the (*D*_0,*I*_, *VCS*_*MRI*_) plane, as it peaks for (*D*_0,*I*_, *VCS*_*MRI*_) around (1.8 µm^2^/ms; 28 µm). Finally, Fig.7G to Fig. 7J show results from the 5-fold cross-validation experiments, by scattering ground truth and predicted tumour volume values for different statistical models. The highest correlation between ground truth and predicted volumes is obtained for a model where *D*_0,*I*_ and *VCS*_*MRI*_ are used as predictors, i.e., a moderate but significant Pearson’s *r* = 0.399 (*p* = 1.02 ‧ 10^-6^; range of *r* across the 5 folds of (0.143; 0.537)). This surpsasses values obtained when other dMRI metrics are used, e.g., DKI ADC and kurtosis *K* (*r* = 0.240, *p* = 4.25 ‧ 10^-3^; range of *r* across the 5 folds of (0.217; 0.404)). Supplementary Data S3 reports all liver tumour values used for these analyses.

**Fig. 7.**
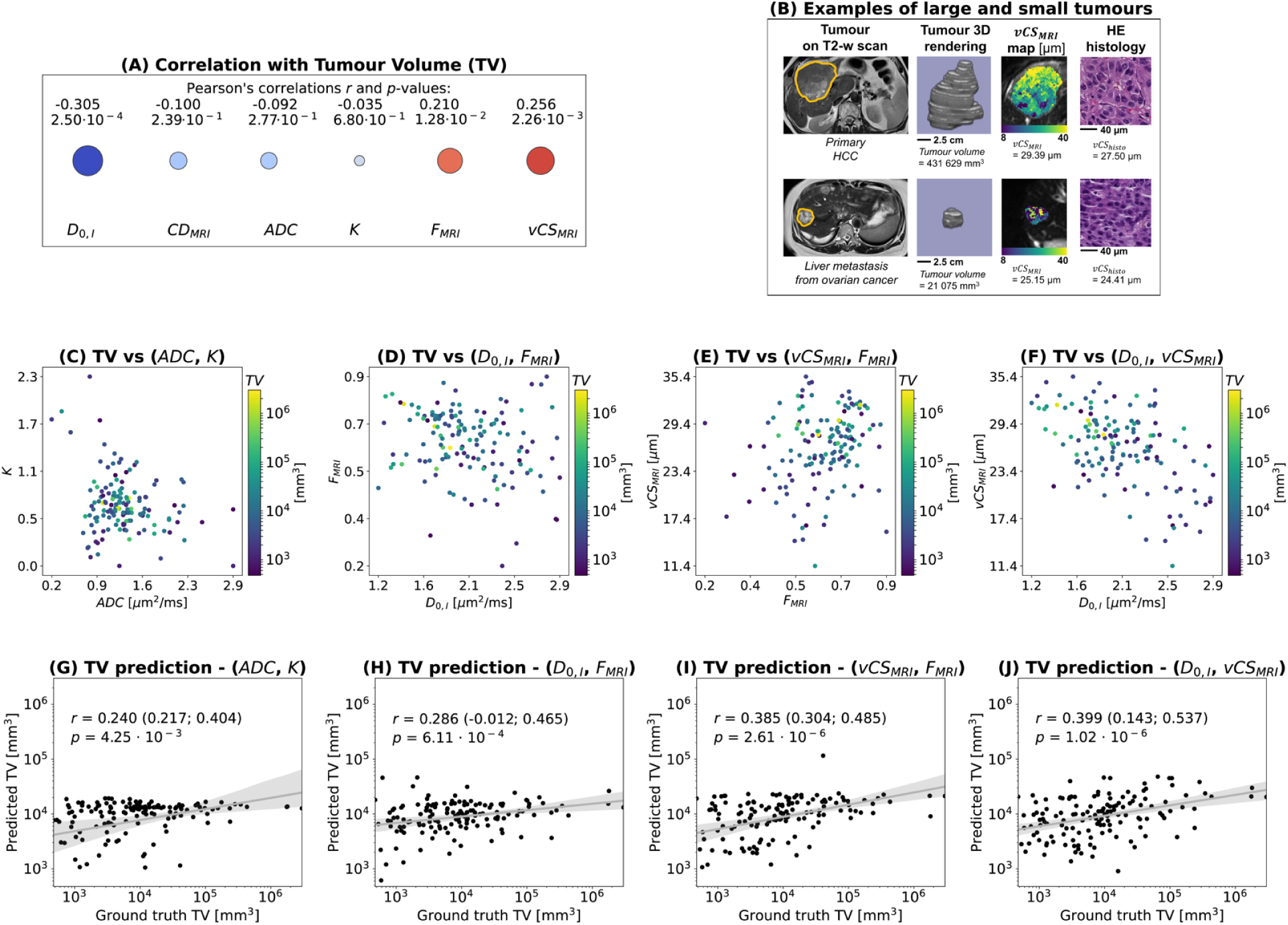
Association between dMRI metrics and liver tumour volume in patients. **(A)**: Pearson’s correlation coefficients between liver tumour volume and per-tumour mean values of *Diff-in* and DKI metrics. **(B)**: examples of large and small tumours from a primary hepatocellular carcinoma (large) and a metastatic ovarian cancer (small), with corresponding *VCS*_*MRI*_ dMRI metric of cell size and HE histology. **(C) to (E)**: visualisation of tumour volume as a bi-dimensional function of different pairs of dMRI metrics, namely: DKI ADC and *K* in **(C)**; *Diff-in D*_0,*I*_and *F*_*MRI*_in **(D)**; *Diff-in VCS*_*MRI*_and *F*_*MRI*_in **(E)**; *Diff-in D*_0,*I*_ and *VCS*_*MRI*_in **(F)**. **(G) to (J)**: prediction of tumour volume based on different pairs of input dMRI, in 5-fold cross validation experiments. These were: DKI ADC and *K* in **(G)**; *Diff-in D*_0,*I*_ and *F*_*MRI*_ in **(H)**; *Diff-in VCS*_*MRI*_and *F*_*MRI*_in **(I)**; *Diff-in D*_0,*I*_and *VCS*_*MRI*_in **(J)**. In panels (G) to (J), scatter plots of ground truth tumour volumes against predicted tumour volumes are obtained by pooling together predictions from all 5 cross-validation folds. The figures also report the Pearson’s correlation coefficient between ground truth and predicted tumour volume values, the linear regression line with 95% confidence intervals, and the range of correlation coefficients obtained across cross-validation folds. Note that in these experiments, the log_10_ of the tumour volume was studied, since the tumour volume spans several orders of magnitude and is therefore challenging to handle numerically.

### The proposed dMRI model distinguishes colorectal cancer from melanoma liver metastases

Lastly, we tested whether the proposed dMRI approach may complement standard diffusion metrics, such as *ADC*, to aid the interpretation of between-tumour differences. For this purpose, we compared the composition of liver metastases from colorectal cancer (CRC) and melanoma, the two most frequent primary cancers in our cohort. We tested for differences in dMRI metrics between the two groups, verifying results with histology. These two cancers feature distinct cellular composition: while CRC metastases are characterised by the presence of large luminal spaces, melanoma metastases consist of tightly packed malignant melanocytes (Fig. 8, panels A and B)^40,41^. The proposed *Diff-in* model provides estimates of IC fraction *F*_*MRI*_ that are significantly larger in melanoma than in CRC metastases (*p* = 0.011, N = 21; Fig. 8, panels A, B and K), a finding that is consistent with the presence of EC water trapped within CRC luminal spaces. This result is confirmed by the histological IC fraction *F*_*histo*_, which is also lower in CRC than in melanoma (*p* = 0.034, N = 12; Fig. 8, panels A, B and E). More standard diffusion indices also detects differences between the two cancer types: DKI *ADC* is higher in CRC than in melanoma metastases (*p* = 0.038, N = 21; Fig. 8, panels A, B and I), also consistent with presence of free diffusion in the lumina. No statistically significant differences between CRC and melanoma were detected for any other metric, including patient’s age.

**Fig. 8.**
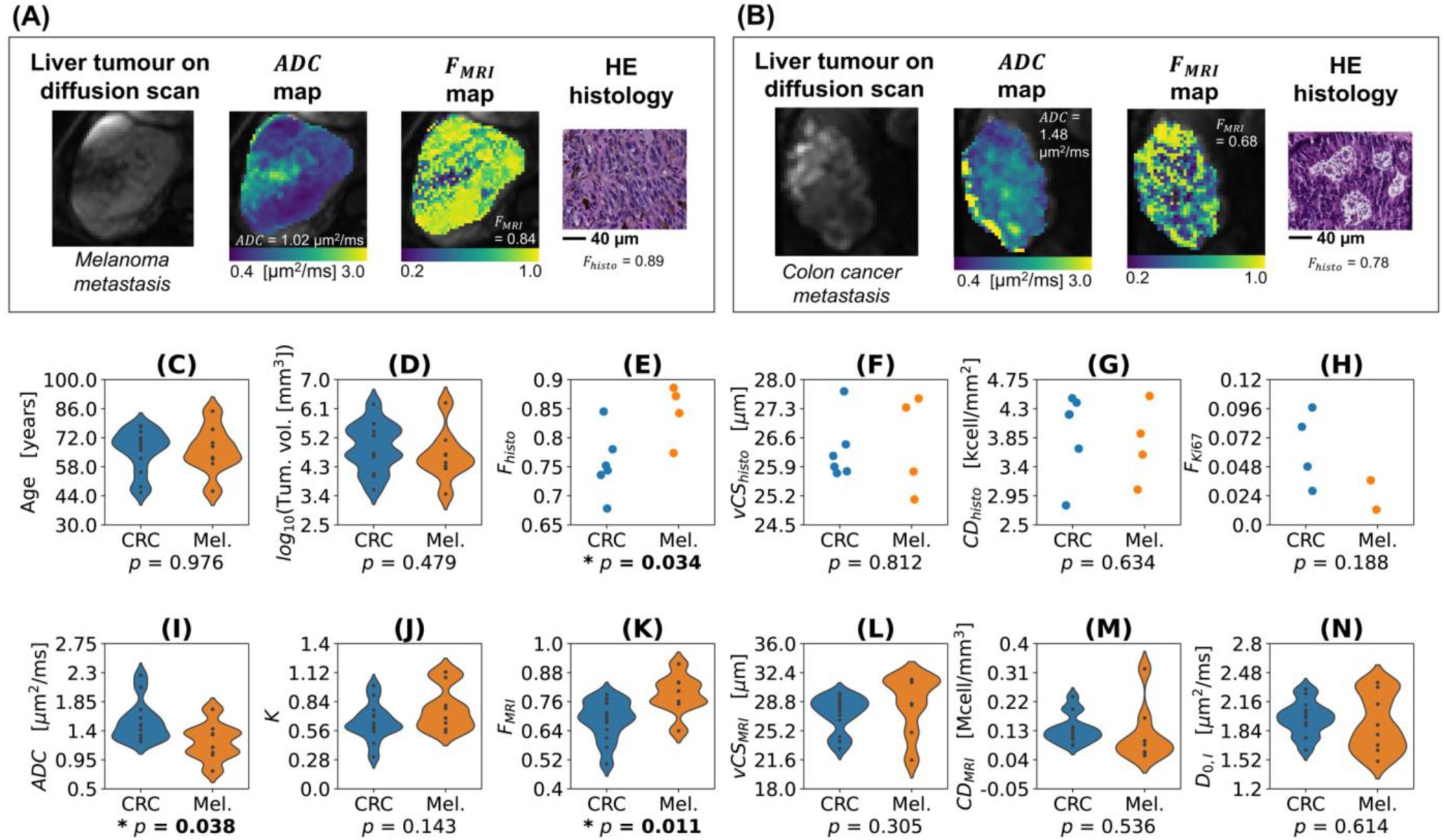
Differences in dMRI and histological metrics between liver metastases from melanoma and colorectal cancer (CRC). Visualisation of illustrative parametric maps in a liver metastasis from melanoma and CRC, and differences between the two groups across dMRI and histological metrics. **(A)**: data from a melanoma liver metastasis (from left to right: DW image, ADC map and *F*_*MRI*_map from dMRI; HE histology, with quantitative IC fraction *F*_*histo*_). **(B)**: same data as in (A), but for a colon cancer liver metastasis. **(C):** age differences between the CRC and melanoma groups (N = 13 CRC; N = 8 melanoma). **(D)**: tumour volume differences (N = 13 CRC; N = 8 melanoma). **(E):** histological IC fraction *F*_*histo*_ differences (N = 6 CRC; N = 4 melanoma). **(F)**: histological cell size *VCS*_*histo*_ differences (N = 6 CRC; N = 4 melanoma). **(G)**: histological cell density *CD*_*histo*_differences (N = 6 CRC; N = 4 melanoma). (H): histological Ki67 staining fraction *F*_*Ki*67_ differences (N = 4 CRC; N = 2 melanoma). **(I)**: DKI *ADC* differences (N = 13 CRC; N = 8 melanoma). **(J)**: DKI kurtosis *K* differences (N = 13 CRC; N = 8 melanoma). (K): *Diff-in* model IC fraction *F*_*MRI*_differences (N = 13 CRC; N = 8 melanoma). **(L)**: *Diff-in* model cell size *VCS*_*MRI*_ differences (N = 13 CRC; N = 8 melanoma). **(M)**: *Diff-in* model cell density *CD*_*MRI*_ differences (N = 13 CRC; N = 8 melanoma). **(N)**: *Diff-in* model IC diffusivity *D*_0,*I*_ differences (N = 13 CRC; N = 8 melanoma).

## Discussion

The latest liver dMRI models aim to disentangle IC and EC water contributions to the total signal^6,17,20,21^. This powerful approach enables the estimation of innovative tissue property maps, but its clinical deployment is hampered by the high number of unknown tissue parameters to estimate, which requires impractically long dMRI acquisitions^17,42,43^. With this challenge in mind, this paper delivers a practical implementation of a two-compartment dMRI signal model, tailored for liver imaging, and truly feasible in the clinic. Through histology-informed model selection, we design a compact framework consisting of fitting a model of restricted IC diffusion to highly DW images, with negligible EC signal contributions. This provides cell size and density estimates that correlate with histology, and which may provide additional information to standard-of-care imaging for non-invasive tumour phenotyping.

To find the optimal dMRI signal implementation, we analysed co-localised dMRI and histology data (N = 25) from fixed mouse livers and from human tumours of the liver. We compared 5 signal models, each fitted according to two distinct strategies, and ranked them for their ability to estimate histological IC fraction and cell size (the uknown tissue parameters in the dMRI signal models). Rankings unequivocally suggest the highest radiological-histological agreement is obtained by fitting a model of restricted diffusion within spherical cells, with negligible EC signal contributions – a model here referred to as *Diff-in* –, to images acquired with b-values higher than approximately 900 s/mm^2^ *in vivo* and 1800 s/mm^2^ *ex vivo*. Interestingly, our central result, confirmed by Monte Carlo computer simulations, points towards the use of simpler models of diffusion if these are deployed in appropriate measurement regimes, and is consistent with recent estimates of the EC liver ADC, as high as 2.5 µm^2^/ms^17^. Such a high *ADC*_*E*_implies that the EC signal would decay to roughly 5% or less of its non-DW value even for b-values of 1200 s/mm^2^ (*exp*(–*b ADC_E_*) ≈ 0.05 for *b* = 1.2 ms/µm^2^ and *ADC_E_* = 2.5 µm^2^ ms^-1^), justifying the hypothesis of negligible EC signal contributions^7,39,44^.

Our proposed dMRI modelling approach provides metrics that visualise tissue heterogeneity over a wide field of view, beyond what can be routinely sampled in the clinic with biopsies. *Diff-in* metrics point, for example, to areas of high cell densities in some of the fixed mouse livers, or areas of necrosis within the core of large tumours in patients *in vivo* – facts that are all confirmed by histology. Notably, spatial trends from the dMRI model *Diff-in* agree with metrics from other candidates dMRI signal implementation, e.g., *Diff-in-exFast*, the model selected when fitting is performed on the whole image data set (Fig. S8 and S11). On the one hand, this gives confidence on the robustness of our signal modelling routines. On the other hand, the noisier appearance of metrics *Diff-in-exFast* compared to *Diff-in* is consistent with ill-posedness of complex dMRI models, when they include several unknowns to estimate^39^.

We investigated the potential clinical utility of the proposed *Diff-in* dMRI approach by testing whether its metrics can serve as non-invasive biomarkers of biologically relevant processes in cancer. For this, we conducted three analyses, namely: i) we studied the association between *Diff-in* metrics and cell proliferation; ii) we investigated the relationship between *Diff-in* metrics and macroscopic tumour characteristics, as their volume; iii) we tested whether *Diff-in* metrics reveal between-tumour differences that are histologically meaningful. Regarding the comparison with Ki67 immunostains demonstrating cell proliferation, we found that dMRI cell size and density are sensitive to Ki67 stains of cell proliferation, being respectively moderately and strongly correlated with the staining fraction *F*_*Ki*67_. This finding, confirmed by histological cell size and density from HE-stained biopsies, may indicate that tumours featuring higher proliferation levels contain smaller and more densely packed cells. This is in line with previous *in vitro* measurements, as those obtained, for example, in colon carciomas^45^, in which the time required for malignant cell populations to double in number was shorter for small cells with large nuclei, compared to larger cells but with small nuclei. The main takeaway of this analysis is that cell morphology assessment through our dMRI model may offer sensitivity to key biological processes of cancer biology, as cell proliferation^46^, which are directly linked to clinical outcomes. The reported association between *in vivo* cell size measurements and Ki67 proliferation is a novel finding. However, we acknowledge that its exact biological implications deserve additional elucidation in larger cohorts, given the small size of our sample.

Regarding the analysis on tumour volume instead, our key result is that metrics from the proposed *Diff-in* approach are associated to tumour volume, albeit only moderately, in a data set of N = 140 liver tumours. This fact allowed us to develop simple regression models capable of predicting tumour volume given characteristic per-tumour dMRI metric values. The best prediction was obtained using cell size *VCS*_*MRI*_and cell diffusivity *D*_0,*I*_– statistical properties of the tumour cell ensemble – as regressors (correlation *r* = 0.399, *p* = 1.02 ‧ 10^-6^ between ground truth and tumour volume predictions), surpassing correlations obtained with DKI ADC and *K*, which only provides *r* = 0.240, *p* = 4.25 ‧ 10^-3^.

Taken as a whole, these findings may suggest that biological properties directly related to the morphology of cancer cells may contribute to explaining, at least in part, the macroscopic phenotype of individual tumours, as for example their size. Therefore, our proposed dMRI approach may offer value to complement standard-of-care radiological assessments, which focus on macroscopic tumour descriptors, providing information that is more closely related to the tumour biology. Nevertheless, we acknowledge again that further validation in larger cohorts is required to confirm these findings, since we were not able to validate the dependence of tumour volume on dMRI metrics histologically, given the limited size of the sample of histological data (Supplementary Table S5). Future work will focus on studying areas of active tumour, since necrosis – common in the large tumours contained in our advanced patients’ cohort – may confound in part the associations, being CS values from both automatic histology processing and dMRI less reliable in highly necrotic areas (see, for example, Fig. 5).

As a final demonstration of the potential utility of our approach, we compared *Diff-in* dMRI metrics across melanoma and CRC liver metastases, being these the two most frequent cancers in our advanced cohort. The analysis showed that *Diff-in* model pins down the exact microstructural property underlying the higher *ADC* seen in CRC, i.e., the lower IC fraction, being *F*_*MRI*_higher in CRC than in mealona, in agreement with *F*_*histo*_. This finding points once again towards the higher biological specificity of the proposed approach against standard dMRI techniques such as DKI: these, while sensitive to tissue cellulear properties, provide surrogate markers that are more difficult to interpret, and that pool several tissue properties into one number^43^.

In our data, *ADC* and *K* do offer value beyond standard-of-care tumour size assessment for cancer characterisation, for example showing correlation with cancer cellularity and cell size, a fact that agrees with known literature^34,43^, and reveal inter-tumour differences, as discussed boave for CRC and melanoma. However, we point out that histology-informed, biophysical dMRI techniques as the one proposed in this article attempt to provide truly quantitative indices, with a direct biological interpretation, and thus overcome the issues related to the interpretaibility and biological specificity of DKI. Contrarily to our proposed metrics, *ADC* and *K* are semi-quantitative and protocol-dependent, and their numerical value can change as function of factors as the dMRI sequence effective diffusion time^43,47^. This may be one of the reason contributing, for example, to the disappointing association between DKI metrics and Ki67 cell proliferation, a fact that confirming the need for developing novel dMRI approaches with high biological specificity for liver tumour imaging – one of the key objectives of this work.

We would like to acknowledge the following potential limitations. Firstly, our sample size is relatively small. This paper provides a first demonstration of the potential utility of metrics from the a practical liver dMRI model, e.g., as markers of cell proliferation or of other biological phenomena that drive tumour phenotypes. Nonetheless, while works proposing related dMRI techniques relied on similar^21,31^ (if not even smaller^17,18^), sample sizes, we acknowledge that our exploratory findings require further confirmation in larger cohorts.

Secondly, we point out that results from any dMRI-histology comparison should always be taken with care. Here we related dMRI metrics obtained *in vivo* to histological indices from biopsies. While we were able to identify the tumours from which the biopsies were taken, we could not identify exactly the tumour area that was biopsied. This implies that a biopsy may not be fully representative of the characteristics of an entire tumour, so that the true extent of the associations between dMRI and histology may have been underestimated. Also, histology has its own limitations *per se*, since it provides readouts that may not be fully accurate. For example, routine HE histology is an inherently 2D technique, unlike 3D MRI. Moreover, it is affected by artifacts (e.g., due to dehydration, paraffin embedding, imperfect staining, cutting, etc^48^), and the automatic processing of large fields-of-view requires trading off between sensitivity and specificity. We took steps to mitigate these issues, e.g., by accounting for biases due to tissue shrinkage. Nonetheless, our histology-derived estimates of cell properties are likely biased versions of the true figures.

We would also like to acknowledge that the proposed dMRI approach neglects other potentially relevant microstructural properties, such as water exchange between intra-/extra-cellular spaces^19,32^, presence of cell size/cytosolic diffusivity distributions^37,49^, or intra-compartmental T2 or T1^31^. On the one hand, ignoring these properties may have biased the estimation of *F*_*MRI*_ and *VCS*_*MRI*_^19,32^. On the other hand, properties such as exchange rates, overlooked in our model, may be relevant markers of cellular stress *per se*. In future, we plan to incorporate these properties in our models, while ensuring the clinical feasibility of the dMRI protocols required to fit them.

To conclude, *this study delivers a practical liver dMRI signal model consisting of restricted diffusion within spherical cells, with negligible EC signals, which should be fitted to b-values higher than, approximately, 900 s/mm^2^* in vivo. This model offers estimates of cell size and density that are correlated to the underlying histology, and which may provide complementary information to routine standard-of-care imaging, as for example for the characterisation of cell proliferation, or for non-invasive tumour phenotyping *in vivo*.

## Methods

### dMRI models

Common biophysical body dMRI signal models^6,17,18,44,50,51^ describe the signal as arising from three, non-exchanging proton pools: vascular water; restricted, intra-cellular water; hindered, extra-cellular, extra-vascular water. The dMRI signal for a PGSE measurement at b-value *b*, gradient duration/separation δ/ Δ, and echo time TE is

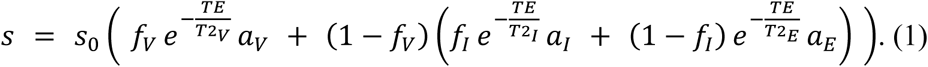

Above, *s*_0_ is the apparent proton density, *f*_*v*_ is the voxel vascular signal fraction, *f*_*I*_ is the tissue intra-cellular signal fraction, *T*2_*v*_/*T*2_*I*_/*T*2_*E*_ and *a*_*v*_/*a*_*I*_/*a*_*E*_ are compartment-wise T2 and diffusion-weighting factors. *a*_*v*_captures intra-voxel incoherent motion (IVIM) effects^52^. *In vivo*, the IVIM vascular ADC ranges approximately^53^ in [15; 60] µm^2^ ms^-1^. For this reason, for *b* > 100 s/mm^2^, the vascular signal vanishes (*a*_*v*_ ≈ 0), and Eq. (1) reduces to^17^

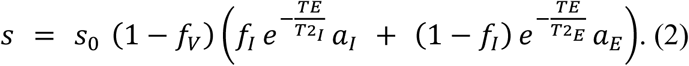

A common model for *a*_*I*_ in Eq. 2 is that of restricted diffusion within spheres of diameter *L*^17,18^:

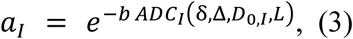

where

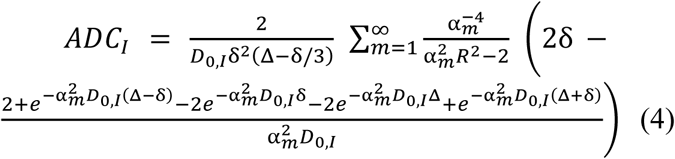

is the Gaussian phase distribution approximation of the intra-cellular ADC^54^. Above, *α_m_* is the *m*-th root of *α_m_R J’_3/2_(α_m_R) – 0.5 J_3/2_(α_m_R) = 0*, *J_3/2_(x)* is the Bessel function of the first kind and order 3/2, and *J’_3/2_(x)* its first-order derivative. *ADC*_*I*_ depends on the intrinsic cytosol diffusivity *D*_0,*I*_ and on the cell size *L* = 2*R* (*R*: radius; *L*: diameter). Noting that dMRI-derived *L* represents a volume-weighted mean cell size statistics^7,37^, we will refer to it as *volume-weighted cell size* (*VCS*).

Conversely, the extra-cellular, extra-vascular signal may be described in terms of hindered diffusion in a tortuous space^17,22,51^:

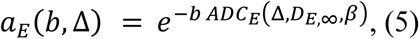

with

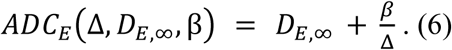

In Eq. 6, *D*_*E*,∞_ is the asymptotic^51^ *ADC*_*E*_ for Δ → ∞.

#### The 5 implementations of the two-compartment model

We investigated 5 implementations of Eq. 2, divided into two families. The first family includes models that do not make assumptions on which of *ADC*_*I*_/*ADC*_*E*_ is higher:

i. *Diff-in-exTD*: the most general model, relying on the full expression of *ADC*_*E*_ in Eq. 6;
ii. *Diff-in-ex*: a simpler implementation of *Diff-in-exTD* that neglects extra-cellular TD (*β* = 0 in Eq. 6).

In the second family of models, we constrain *ADC*_*E*_ > *ADC*_*I*_. It includes

i. *Diff-in-exTDFast*: equivalent to *Diff-in-exTD*, with the lower bound for *D*_*E*,∞_ ensuring *ADC*_*E*_ > *ADC*_*I*_ for any *L*.
ii. *Diff-in-exFast*: equivalent to *Diff-in-ex*, but again ensuring that *ADC*_*E*_ > *ADC*_*I*_ for any *L*.
iii. *Diff-in*: a model where the extra-cellular signal is negligible compared to the intra-cellular one, due to *ADC*_*E*_ being much larger than *ADC*_*I*_, so that Eq. 2 simplifies to

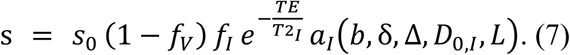

In all models we used *T*2_*I*_ ≈ *T*2_*E*_ ≐ *T*2_*T*_, given the challenge of resolving accurately multiple T2 constants^18,31^.

#### Fitting

We fitted the 5 models using custom-written Python routines, based on objective function minimisation initialised by a grid search. The objective function was *f*_*obj*_ = −*ln*(λ), where λ is the offset-Gaussian likelihood^29^. Fitting provides estimates of *VCS* and voxel IC signal fraction

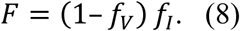

We also combined *VCS* and *F* into a cell density per unit volume^18^

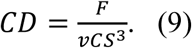

### Mouse data acquisition

#### Animals

We obtained data from 7 fixed livers of NOD.Cg-Prkdc^scid^ IL2rg^tm1WjI^/SzJ mice. All experimental protocols were approved and monitored by the Vall d’Hebron Institute of Research Animal Experimentation Ethics Committee (CEEA; registration number 68/20) in accordance with relevant local and EU regulations. We studied six livers from mice implanted with cells derived from biopsies of prostate cancer patients, as part of an ongoing study, plus an additional liver from a mouse without any implantation. We implanted one tumor biopsy core with growth factor-enriched Matrigel (Corning) subcutaneously in the flank of each mice. We derived tissue from the following biopsies: iliac bone metastasis biopsy (metastatic castration-resistant prostate cancer, presenting with bone metastasis and Gleason score 3+4 adenocarcinoma); prostate biopsy (patient with metastatic hormone-sensitive prostate cancer, presenting with bone metastasis and Gleason score 5+4 adenocarcinoma); two liver biopsies (patient with metastatic castration-resistant prostate cancer, presenting with bone and visceral metastasis and Gleason score 4+4 acinar adenocarcinoma; patient with metastatic hormone-sensitive prostate cancer, presenting with bone and liver metastasis and Gleason score 4+4 adenocarcinoma). After implantation, we measured tumour size using calipers and monitored mouse weight weekly, sacrificing animals by cervical dislocation under general anesthesia when tumour volume exceeded 2000 mm^3^. We collected the livers, fixed them overnight in formalin, and transferred them to phosphate-buffered saline (PBS) solution.

#### MRI

We scanned livers on a 9.4T Bruker Avance system at room temperature. Livers were tightened with sewing thread to a histology cassette and placed into a Falcon^®^ tube, filled with PBS solution. A 1-channel birdcage coil was used (excitation/reception). The protocol included a T2-weighted fast spin echo sequence (resolution: 144 μm × 144 μm × 2.216 mm) and PGSE dMRI (Fig. S13A; TR = 2700 ms; resolution: 386 μm × 386 μm; matrix size: 86 × 86; 4 slices, 2.216 mm-thick, NEX = 1). The protocol featured: δ = 10 ms, Δ = {15, 30} ms, 10 linearly spaced b-values for each Δ (minimum/maximum nominal *b*: 0/2800 s/mm^2^). DW images corresponding to Δ = 15 ms were acquired at each of TE = {31, 45, 65} ms, and to Δ = 30 ms at each of TE = {45, 65} ms. We i) denoised dMRI scans with Marchenko-Pastur Principal Component Analysis (MP-PCA) Python denoising^55^ (kernel: 7×7×3), ii) mitigated Gibbs ringing (MrTrix3 local sub-voxel shift method^56^), and iii) corrected temporal signal drifts by assessing signal changes in a PBS solution region, accounting for TE (PBS T2: 500 ms).

Finally, we fitted the *Diff-in-exTD*, *Diff-in-exTDFast*, *Diff-in-ex*, *Diff-in-exFast* and *Diff-in* models voxel-by voxel (tissue parameter bounds: [0; 1] for *f*_*I*_; [0.8; 2.6] μm^2^ ms^-1^ for *D*_0,*I*_; [8; 40] µm for *vCS*; [0.8; 2.6] μm^2^ ms^-1^ for *D*_*E*,∞_ in models *Diff-in-ex* and *Diff-in-exTD* and [1.75; 2.6] μm^2^ ms^-1^ in models *Diff-in-exFast* and *Diff-in-exTDFast*; [0; 10] μm^2^ for *β* in models *Diff-in-ex-TD* and *Diff-in-exTDFast*). For fitting, we fixed *f*_*v*_ and *T*2_*T*_ to values obtained through a a two-pool vascular-tissue model^57^ (fitting bounds: [0; 1] for *f*_*v*_; [5; 80] ms for *T*2_*T*_). Fitting was performed i) on all images with *b* > 1000 s/mm^2^ (suppressing vascular signals, referred to as *fitting on whole image set*); ii) on *b* > 1000 s/mm^2^ images (*high b-value fitting*). In our *ex vivo* data, the vascular signal captures PBS solution contamination (PBS ADC: roughly 2.4 μm^2^ ms^-1^). For this reason, we adopted a b-value threshold of 1000 s/mm^2^ to achieve acceptable PBS signal suppression. We used instead a minimum b-value of 1800 s/mm^2^ for high b-value fitting (minimising extra-cellular contributions), given the reduction in intrinsic tissue diffusivity expected *ex vivo*.

For comparison, we computed ADC and apparent diffusion excess kurtosis *K* by fitting

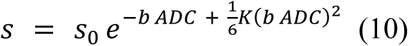

to DW images acquired at *TE* = 45 ms, Δ = 30 ms, with in-house Python code.

#### Histology

After MRI, samples underwent histology. We cut two 4 μm-thick histological sections at known position, stained them with HE, and digitised them (Hamamatsu C9600-12 slide scanner; 0.227 μm resolution). An experienced pathologist (S.S.) inspected images qualitatively. We then processed them with the automatic cell detection tool of QuPath^27^, obtaining per-cell area *A* and diameter *l* = √ 4/π *A*. Afterwards, we split images into 386 μm × 386 μm patches (matching the MRI resolution), computing patch-wise histological volume-weighted cell size *VCS*_*histo*_, intra-cellular area fraction *F*_*histo*_ and cell density per unit area *CD*_*histo*_^37^. *VCS*_*histo*_, defined as

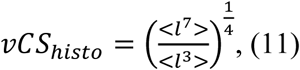

is a more accurate counterpart of dMRI cell size than the arithmetic mean^7,37^ *aCS*_*histo*_ = < *l* >. We accounted for biases coming from: i) estimating the size of 3D objects from 2D views (bias 1), ii) tissue shrinkage (bias 2), by rescaling *VCS*_*histo*_and *CD*_*histo*_. The final *VCS*_*histo*_estimate was 1.4616 times larger than the value obtained from direct image processing (1.4616 = 1.2732×1.148; 1.2732, derived from the theory of spherical caps, accounts for bias 1; 1.148 accounts for bias 2, and corresponds to a plausible shrinkage of 12.9% following dehydration, clearing and paraffin embedding^48^). The final *CD*_*histo*_ estimate was 1.318 times smaller than the value derived from direct image processing, since 1 mm^2^ of shrunk tissue corresponds to 1.148×1.148 mm^2^ = 1.318 mm^2^ of unprocessed tissue (plausible shrinkage 12.9%^48^). Lastly, we co-registered histological maps to MRI^37^ using DiPy^58^.

### Human data acquisition

#### Cohort

We obtained data from patients suffering from advanced solid tumours, recruited for an ongoing imaging study approved by the Vall d’Hebron University Hospital Ethics committee (PR(AG)29/2020). Patients provided informed written consent to participate in the imaging study. We included 38 patients with liver malignancies, who were scanned with either a 1.5T or 3T system. Approximately one week after MRI, a biopsy from one of the imaged liver tumours was obtained from 18 patients. The histological material was stained for HE, and in 12 cases also through Ki67 IHC, demonstrating cell proliferation.

#### MRI

We imaged patients at the level of the abdomen. We scanned 11 patients on a 1.5T Siemens Avanto scanner using the vendor 18-channel body coil for detection, and 27 patients on a 3T GE SIGNA Pioneer scanner, using the vendor 48-channel torso coil for signal reception, with 32 channels enabled for detection.

In the 1.5T Siemens system, the protocol included a T2-weighted fast spin echo scan (resolution: 1.4 × 1.4 × 5 mm^3^; 32 slices; TR = 4500 ms; TE = 82 ms; echo train length: 29; NEX = 8; GRAPPA = 2) and fat-suppressed DW TRSE (Fig. S13B) EPI (dMRI scan time: 16 minutes). It featured: resolution: 1.9 × 1.9 × 6 mm^3^; 32 slices; TR = 7900 ms; bandwidth 1430 Hz/pixel; averaging of 3 orthogonal diffusion directions × 2 signal averages (effective NEX = 6); GRAPPA factor of 2; 6/8 partial Fourier imaging. The dMRI protocol consisted of *b* = {0, 50, 100, 400, 900, 1200, 1600} s/mm^2^, each for TE = {93, 105, 120} ms. One additional image (*b* = 0 s/mm^2^; TE = 93 ms) was acquired with reversed phase encoding polarity. The gradient timings (Fig. S13B) were: δ_1_ = 8.9 ms, δ_2_ = 17.6 ms, δ_3_ = 20.4 ms, δ_4_ = 6.0 ms, Δ_1,2_ = 17.4 ms and Δ_1,4_ = 63.9 ms when TE = 93 ms; δ_1_ = 13.2 ms, δ_2_ = 19.3 ms, δ_3_ = 24.8 ms, δ_4_ = 7.7 ms, Δ_1,2_ = 21.7 ms and Δ_1,4_ = 74.2 ms when TE = 105 ms; δ_1_ = 18.9 ms, δ_2_ = 21.0 ms, δ_3_ = 30.5 ms, δ_4_ = 9.5 ms, Δ_1,2_ = 27.5 ms and Δ_1,4_ = 87.5 ms when TE = 120 ms. The b-value is

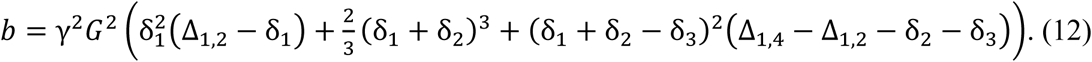

Converely, the protocol implemented on the 3T GE scanner included a respiratory-gated T2-weighted fast spin echo scan (resolution: 1.4 × 1.4 × 6 mm^3^; 32 slices; TR = 4615 ms; TE = 52.86 ms; echo train length: 16) and respiratory-gated, fat-suppressed PGSE (Fig. S13A) EPI (dMRI scan time: 16 minutes). It featured: resolution: 2.4 × 2.4 × 6 mm^3^; 32 slices; TR = 6000 ms; bandwidth 1953 Hz/pixel; averaging of 3 orthogonal diffusion directions × 2 signal averages (effective NEX = 6); ASSET factor of 2. The dMRI protocol consisted of *b* = {0, 50, 100, 400, 900, 1200, 1500} s/mm^2^, each for TE = {75, 90, 105} ms. The gradient timings (Fig. S17A) were: gradient duration δ = {0.0, 3.9, 5.2, 9.2, 15.0, 18.2, 21.0} ms for TE = 75 ms, δ = {0.0, 3.9, 5.2, 9.2, 13.0, 15.8, 18.5} ms for TE = 90 ms and 105 ms; gradient separation Δ = {0.0, 27.8, 29.0, 33.0, 28.7, 31.8, 34.7} ms for TE = 75 ms and Δ = {0.0, 27.8, 29.0, 33.0, 37.0, 39.6, 42.3} ms for TE = 90 ms and TE = 105 ms.

dMRI post-processing consisted of slice-wise Python MP-PCA denoising (kernel: 5×5)^55^; MRTrix3 Gibbs unringing^56^; motion correction via affine co-registration^59^; FSL distortion correction^60^ (1.5T data only). An experienced radiologists (R.P.L.) segmented tumours on the T2-w scan, enabling per-patient tumour volume computation. Afterwards, we warped the tumour mask to dMRI using ANTs^61^ non-linear co-registration, and fitted the 5 dMRI models, fixing again *f*_*v*_ and *T*2_*T*_to previously computed values^57^ (fitting bounds: [0; 1] for *f*_*v*_; [20; 140] ms for *T*2_*T*_; [0; 1] for *f*_*I*_; [0.8; 3.0] μm^2^ ms^-1^ for *D*_0,*I*_; [8; 40] µm for *vCS*; [0.8; 3.0] μm^2^ ms^-1^ for *D*_*E*,∞_in models *Diff-in-ex* and *Diff-in-exTD,* and [1.75; 3.0] μm^2^ ms^-1^ in models *Diff-in-exFast* and *Diff-in-exTDFast*; [0; 10] μm^2^ for *β* in models *Diff-in-ex-TD* and *Diff-in-exTDFast*).

We fitted the 5 dMRI models i) on images acquired at a b-value *b* > 100 s/mm^2^, to suppress vascular signals (*fitting to the whole image set*); ii) to *b* > 900 s/mm^2^ images, to also minimize extra-cellular contributions (*high b-value fitting*). For scans performed on the 1.5T Siemens system: i) we used Δ_1,2_ + δ_2_ in place of Δ in Eq. 6 (Fig. S13B), ii) we replaced Eq. 4 with a numerical implementation of restricted diffusion within spheres, based on Radial Basis Function interpolation of synthetic signals generated for DW-TRSE with Monte Carlo simulations^62^.

For both scanners, we also computed ADC and excess kurtosis *K* by fitting Eq. 10 on *b* > 100 s/mm^2^ images (shortest TE), with in-house Python code.

#### Histology

We performed ultrasound-guided biopsies of one liver tumour at the Barcelona Vall d’Hebron University Hospital (Spain). Biopsies were obtained approximately one week after dMRI. In two patients, the biopsy was obtained after receiving immunotherapy as part of a phase I trial. In those two cases, an additional dMRI scan was also acquired after starting treatment, immediately before the biopsy. The biological material underwent standard processing, HE staining and Ki67 IHC, and final digitalisation on a Hamamatsu C9600-12 slide scanner (resolution: 0.454 μm). An experienced pathologist (S.S.) assessed the images and drew a region-of-interest (ROI) outlining tumour tissue and excluding non-tumour meterial. In parallel, an experienced radiologist (R.P.L.) inspected ultrasound and MR images, outlining the biopsied tumour on the latter. We processed HE data with QuPath and computed per-biopsy *VCS*_*histo*_, *F*_*histo*_ and *CD*_*histo*_ as previously described. Additionally, we also computed the fraction of tumour labelled for Ki67 (referred to as *F*_*Ki*67_) with in-house routines, in those cases where Ki67 IHC was available (IHC images acquired with the same Hamamatsu scanner used for HE). *VCS*_*histo*_ and *CD*_*histo*_ were rescaled to account for tissue shrinkage. *VCS*_*histo*_ was multiplied by 1.503, where 1.503 = 1.1806×1.2732 accounts for two scaling factors, namely: i) 1.2732 accounts for cell size underestimation due to 2D sectioning, ii) 1.1806 accounts for a plausible tissue shrinkage of 15.3% following fixation, dehydration, clearing and paraffin embedding^48^. The final *CD*_*histo*_ estimate was instead 1.3938 times smaller than the value derived from direct image processing, since 1 mm^2^ of shrunk tissue corresponds to 1.1806×1.1806 mm^2^ = 1.3938 mm^2^ of unprocessed tissue for a shrinkage factor of 15.3%^48^.

### Analyses

#### dMRI model selection

We carried out model selection independently for each of the two fitting strategies. The MRI-histology *Total Correlation Score* (TCS) selects the model providing the highest Pearson’s correlation between *VCS*_*MRI*_ and *VCS*_*histo*_, and between *F*_*MRI*_ and *F*_*histo*_. It is defined as

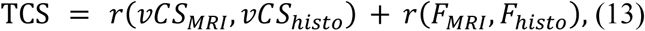

where *r*(*VCS*_*MRI*_, *VCS*_*histo*_) and *r*(*F*_*MRI*_, *F*_*histo*_) are the correlation coefficients of *VCS*_*MRI*_ and *F*_*MRI*_with histological *VCS*_*histo*_and *F*_*histo*_. The correlation between *CD*_*MRI*_and *CD*_*histo*_was not included in Eq. 13 since *CD*_*MRI*_ is determined analytically from *VCS*_*MRI*_ and *F*_*MRI*_. For TCS computation, we pooled together mouse and human data (N = 25, including the two cases who had their biopsy collected after starting immunotherapy).

We also performed model selection using a *Histology Fidelity Criterion* (HFC), and popular *Bayesian Information Criterion* (BIC)^28,29^. HFC rewards the models providing the best accuracy in the numerical estimation of histological cell size and intra-cellular fraction estimation, i.e., minimising

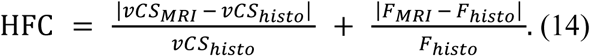

Information on *CD*_*MRI*_ and *CD*_*histo*_ was not included in Eq. 14 since *CD*_*MRI*_ is not a degree of freedom of the dMRI models (it is determined analytically from *VCS*_*MRI*_and *F*_*MRI*_). BIC selects the model providing the best goodness of fit, penalising complexity, by minimising λ is

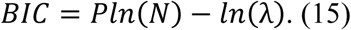

the maximised likelihood, and *P*/*N* are the number of model parameters/signal measurements. We performed BIC selection voxel-wise, followed by majority voting across voxels.

#### Simulated dMRI model selection

We validated results from MRI and histology data through computer simulations. For this experiment, we synthesised signals for each of the three dMRI protocols considered in the study, fitted the 5 candidate dMRI models, and performed model selection using TCS, HFC and BIC. dMRI signals were synthesised through Monte Carlo simulations, which we performed using the MCDC simulator^62^. We seeded walkers in a substrate made of spherical cells of identical diameter^6,17,20,21^ (Fig. S4), controlling the intra-sphere fraction *F* by adding gaps of increasing size in-between abutting spheres, packed in an ideal cubic lattice. We probed four *F* values (0.197, 0.323, 0.406, 0.523) and four sphere diameters for each *F* (8, 16, 22 and 30 µm). We varied intra-/extra-sphere diffusivities (10×10 values; [0.8; 2.6] µm^2^ ms^-1^ for the *ex vivo* protocol and [0.8; 3.0] µm^2^ ms^-1^ for *in vivo* protocols), for a total of 1600 synthetic voxels. We corrupted synthetic signals with Rician noise (*b* = 0 signal-to-noise ratio: 30), and performed model selection according to TCS, HFC and BIC.

#### dMRI-histology correlation

We computed mean and standard deviation of all metrics i) within the mouse liver samples, ii) within a mask containing all liver tumours in patients, iii) within the biopsied patients’ tumours. We pooled together metrics from mice and patients to calculate Pearson’s correlation coefficients *r*, as described above. ADC was normalised to the ADC of the PBS solutions in mice and to the free water diffusivity in patients (3.0 μm^2^ ms^-1^ at 37 °C), given the difference in temperature.

#### Association with immunohistochemical markers of cell proliferation

We assessed the association between *F*_*Ki*67_, an IHC index assessing the fraction of biopsied tissue stained for Ki67 and demonstrating cell proliferation, with dMRI and HE-derived histological properties. For this purpose, we calculated the Pearson’s correlation coefficient between *F*_*Ki*67_ and all dMRI and HE-derived metrics. We excluded two cases whose biopsies were obtained after having started receiving immunotherapy as part of a phase I trial.

#### Association with liver tumour phenotype

Finally, we tested whether metrics from the proposed dMRI approach contribute to explaining the macroscopic tumour phenotype. To this end, we studied the association between dMRI metrics and tumour volume, and compared their values, alongside histological metricsand patients’ age, between CRC and melanoma liver metastases through t-tests. Before performing these comparisons, we harmonised dMRI metrics across scanners with a custom-written ComBat^63^ implementation, rescaling metrics obtained on the 1.5T system to the 3T range. This was performed to account for the fact that certain tumour types (e.g., CRC) were scanned predominantely with one MRI system. Both tumour volume assessment and CRC-melanoma comparison were performed also for histological metrics, excluding two cases who had entered an immunotherapy phase I trial, and whose biopsy was obtained after receiving treatment.

Regarding the tumour volume assessment, we computed Pearson’s correlation coefficients between mean values of dMRI metrics within each individual liver tumour and the tumour volume (N = 140 liver tumours from 38 patients; 137 metastases, 3 primary cancers). Moreover, we performed experiments in which we attempted to predict liver tumour volume given per-tumour mean values of dMRI metrics. Following a standard 5-fold cross validation design, we split the set of tumours into training and test sets. Afterwards, we used the training set to fit linear statistical models of the form

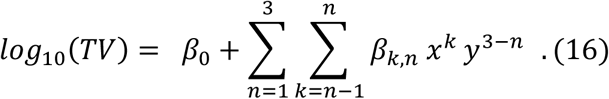

Above, *Tv* is the tumour volume in mm^3^, modelled as a 3^rd^ order polynomial function of *x* and *y*, which is linear in the (unknown) β coefficients of the polynomial terms. *x* and *y* in Eq. 16 represent in turn different pairs of dMRI metrics, as for example DKI *ADC* and *K*, or pairs of metrics from the *Diff-in* model (we tested explicitly *VCS*_*MRI*_ and *F*_*MRI*_; *D*_0,*I*_ and *F*_*MRI*_; and *D*_0,*I*_ and *VCS*_*MRI*_). Afterwards, we deployed the trained statistical models on the test data, calculating the Pearon’s correlation coefficient between ground truth and predicted *log*_10_(*Tv*) values, pooling together predictions from all 5 cross-validation folds. Note that the log_10_ of the volume was studied instead of the volume itself, since the latter spans almost 4 orders of magnitude in our data set, and is therefore difficult to handle numerically.

## Author contributions

**Conceptualization:** F.G., R.P.L., K.B., M.P., E.G., R.T., P.N., J.M. **Methodology:** F.G., R.P.L., K.B., M.P., A.G, D.N.G. **Investigation:** F.G., R.P.L., K.B., I.C.S., I.B., S.S., G.S., A.G., D.N.G., C.M., V.G., J.F.C., X.M., R.M., N.R., M.E., M.Vie., R.T., P.N., J.M., E.G. **Resources:** R.P.L., F.G., P.N., J.M., E.G., N.R., M.E., V.G., M.Vid., P.G.P.G, I.B. **Formal analysis:** F.G. **Visualization:** F.G. **Validation:** F.G. **Software:** F.G., K.B., A.G., D.N.G., C.M.. **Data curation:** F.G., R.P.L., K.B., A.V., G.S., I.C.S., A.G, D.N.G.. **Project administration:** F.G., R.P.L., K.B., E.G., P.N., R.T., J.M., I.C.S. **Funding acquisition:** R.P.L., E.G., R.T., P.N., J.M., F.G., K.B., I.C.S. **Supervision:** F.G., R.P.L., E.G., R.T., P.N., J.M. **Writing—original draft:** F.G., R.P.L., K.B., M.P. **Writing—review & editing:** all authors.

## Ethics

All experimental protocols in animals were approved and monitored by the Vall d’Hebron Institute of Research Animal Experimentation Ethics Committee (CEEA; registration number 68/20) in accordance with relevant local and EU regulations. The imaging study in cancer patients was approved by the Vall d’Hebron University Hospital Ethics committee (PR(AG)29/2020), Barcelona, Spain. Patients provided informed written consent to participate in the study.

## Data Availability

Raw MRI and histological images from the mouse livers will be released freely online through the Radiomics Group web site following publication in a peer-reviewed journal. Raw MRI and histological images of patients cannot be made freely available due to ethical restrictions at this stage. Python routines enabling the computation of the diffusion MRI metrics presented in this article will be released freely online after publication in a peer-reviewed journal (https://github.com/fragrussu/bodymritools)

## Acknowledgments

We thank the whole medical oncology, radiology, pathology, molecular biology, clinical trial, and IT teams at the Vall d’Hebron University Hospital and at the Vall d’Hebron Institute of Oncology in Barcelona (Spain), without whom this study would not have been possible. We are also thankful to the Vall d’Hebron Radiology department and to the ASCIRES CETIR clinical team for their assistance, and to past and present members of the Radiomics group for useful discussion and advice. Finally, we would like to express our sincere gratitude to all patients and their families for dedicating their time to research. VHIO would like to acknowledge: the State Agency for Research (Agencia Estatal de Investigación) for the financial support as a Center of Excellence Severo Ochoa (CEX2020-001024-S/AEI/10.13039/501100011033), the Cellex Foundation for providing research facilities and equipment and the CERCA Programme from the Generalitat de Catalunya for their support on this research. This study has been funded by Instituto de Salud Carlos III (ISCIII) through the project “PI21/01019” and co-funded by the European Union. Data acquisition was also supported by PREdICT, sponsored by AstraZeneca. This study has been co-funded by the European Regional Development Fund/European Social Fund ‘A way to make Europe’ (to R.P.L.), and by the Comprehensive Program of Cancer Immunotherapy & Immunology (CAIMI), funded by the Banco Bilbao Vizcaya Argentaria Foundation Foundation (FBBVA, grant 89/2017). R.P.L is supported by the “la Caixa” Foundation CaixaResearch Advanced Oncology Research Program, the Prostate Cancer Foundation (18YOUN19), ISCIII (PI18/01395), a CRIS Foundation Talent Award (TALENT19-05), the FERO Foundation through the XVIII Fero Fellowship for Oncological Research, the Asociación Española Contra el Cancer (AECC) (PRYCO211023SERR) and the Generalitat de Catalunya Agency for Management of University and Research Grants of Catalonia (AGAUR) (2023PROD00178). The project that gave rise to these results received the support of a fellowship from “la Caixa” Foundation (ID 100010434). The fellowship code is “LCF/BQ/PR22/11920010” (funding F.G. and A.V.) and “LCF/BQ/PI20/11760033” (funding I.C.S). I.C.S. also receives the support of the European Union’s Horizon 2020 research and innovation programme under the Marie Sklodowska-Curie grant agreement No 847648. This research has received support from the Beatriu de Pinós Postdoctoral Program from the Secretariat of Universities and Research of the Department of Business and Knowledge of the Government of Catalonia, and the support from the Marie Sklodowska-Curie COFUND program (BP3, contract number 801370; reference 2019 BP 00182) of the H2020 program (to K.B.). M.P. is supported by the UKRI Future Leaders Fellowship MR/T020296/2. A.G. is supported by a Severo Ochoa PhD fellowship (PRE2022-102586). C.M. is funded by the Asociación Española Contra el Cancer (AECC) (PRYCO211023SERR).

## Author information

Francesco Grussu and Raquel Perez-Lopez are joint corresponding authors.

---

**Figure S1.**
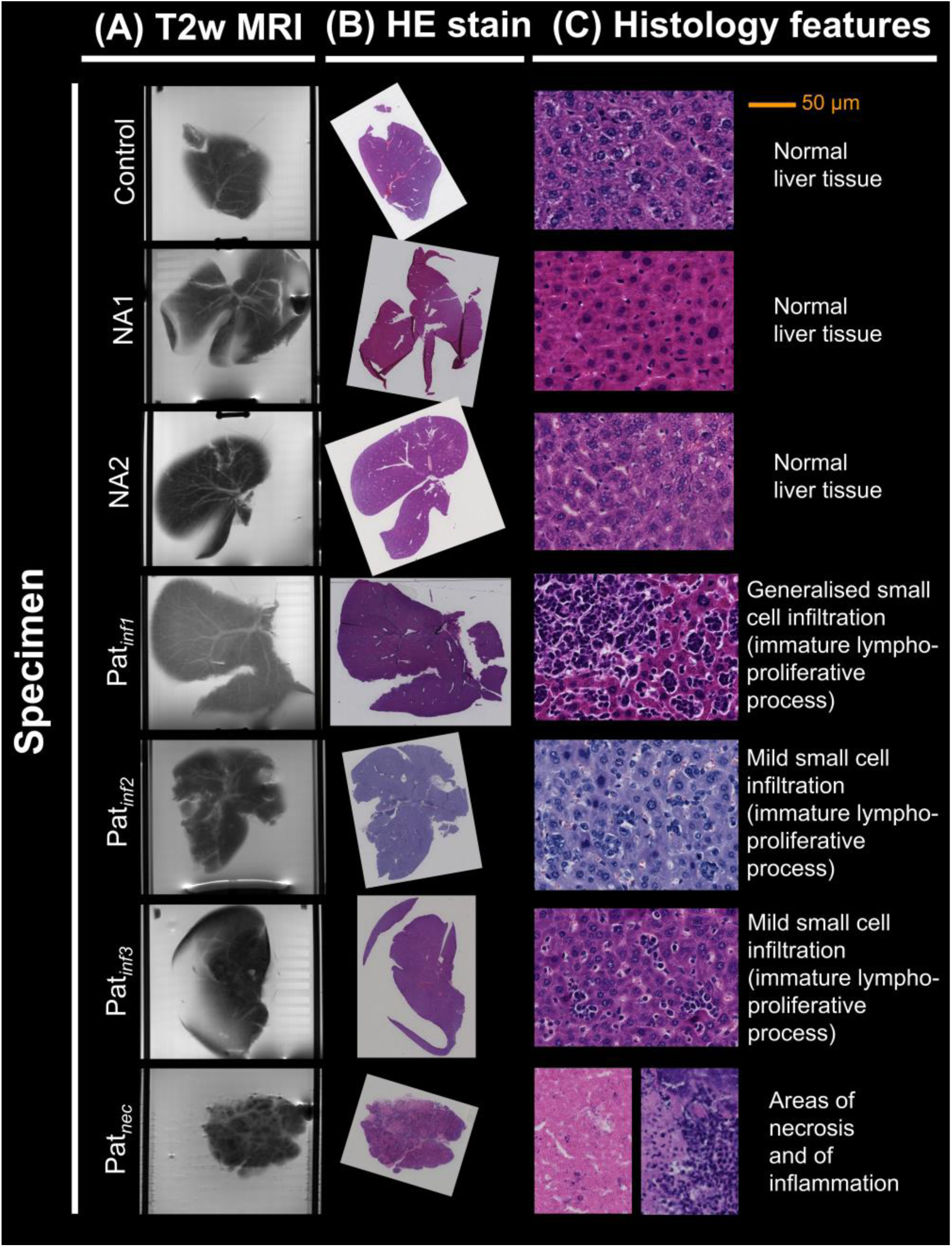
Radiological-histological co-localisation of the *ex vivo* mouse liver data. Illustration of the radiological-histological co-localisation on the 7 fixed mouse livers obtained from mice implanted with a biopsy from a prostate cancer patient. **(A)**, left: illustrative slice of the high-resolution anatomical T2-weighted fast spin echo. **(B)**, centre: hematoxylin and eosin (HE)- stained section, taken from the MRI slice shown to the left. **(C)**, right: detail of the microstructure characterising each specimen, as assessed by an experienced pathologist (SS). Different specimens are arranged along different rows. From top to bottom: *Control*, normal liver structures (no biopsy implantation); *Pat_NA1_* and *Pat_NA2_*, normal appearing normal liver structures after prostate cancer biopsy implantation; *Pat_inf1_*, *Pat_inf2_* and *Pat_inf3_*: pathology following implantation, consisting of an immature, lympho-proliferative process (infiltration of small cells in sinusoidal spaces); *Pat_nec_*, pathology following implantation, consisting of necrosis and inflammation.

**Figure S2.**
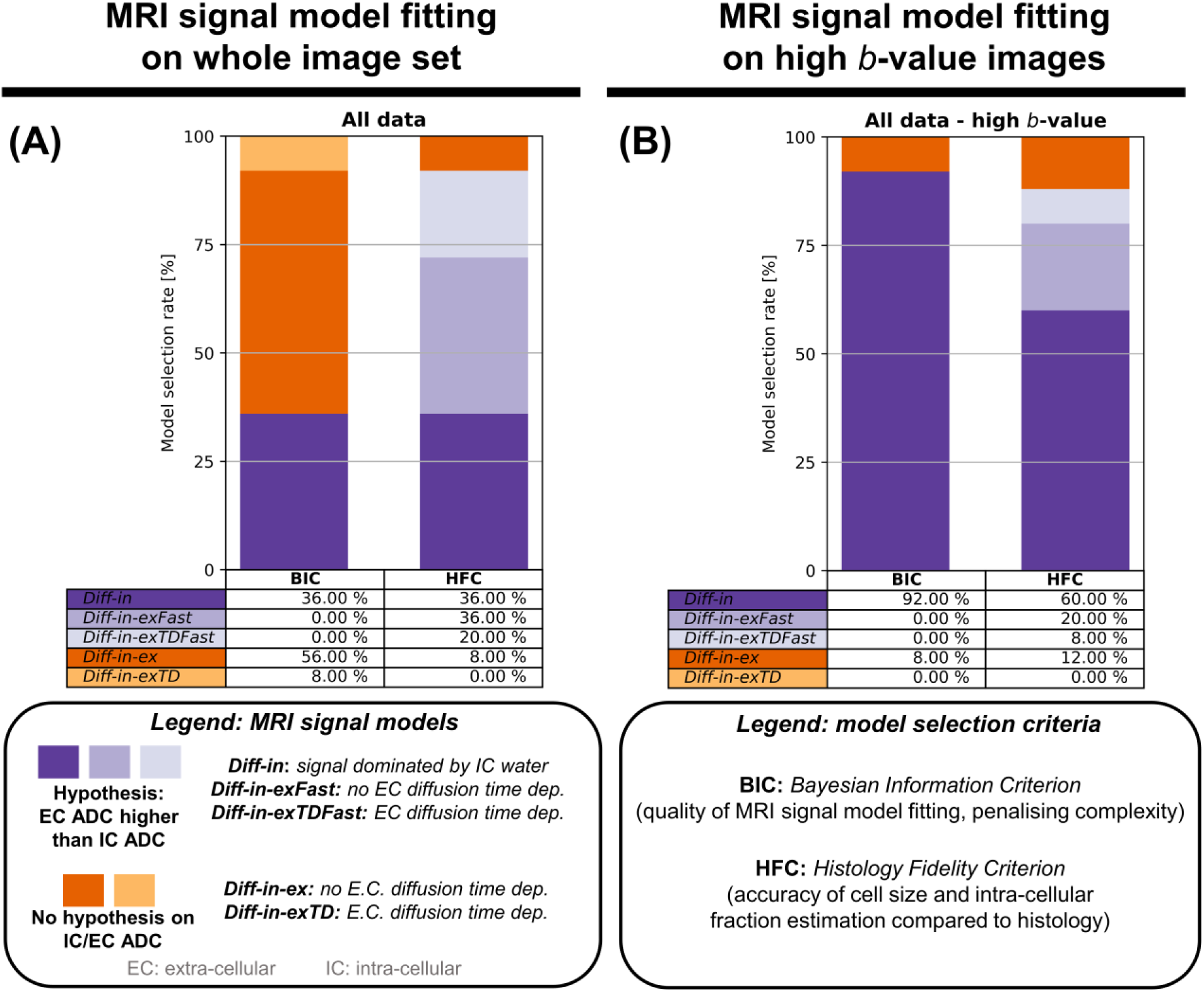
Biophysical dMRI signal model selection based on BIC and HFC criteria. Frequency of model selection based on the *Bayesian Information Criterion* (BIC, quantifying how well a model fits the dMRI signals, penalising model complexity) and on the *Histology-fidelity Criterion* (HFC, quantifying how accurately a dMRI models estimates the intra-cellular fraction and the volume-weighted cell size as seen on histology). (**A**) reports results when models are fitted to the entire set of measurements with negligible vascular constributions (*b* > 1000 s/mm^2^ for suppression of PBS fluid within vessels in the fixed mouse livers; *b* > 100 s/mm^2^ for IVIM signal suppression *in vivo* on clinical systems), while (**B**) reporting results obtained when fitting models only on high b-value images (*b* > 1800 s/mm^2^ in fixed mouse livers; *b* > 900 s/mm^2^ *in vivo*).

**Figure S3.**
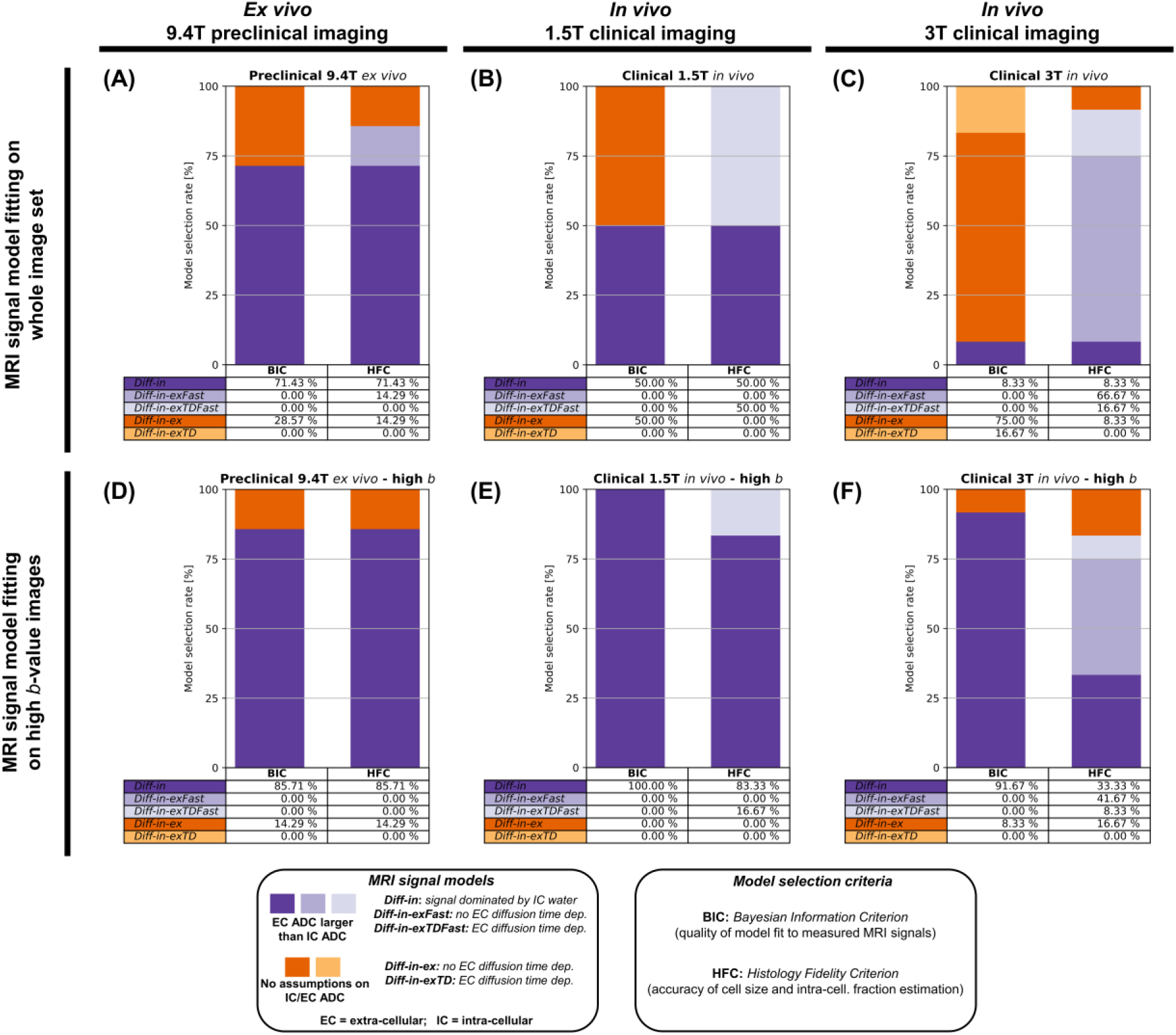
Biophysical model selection with BIC and HFC criteria across different MRI scanners and data subsets. Frequency of selection of each of 5 biophysical dMRI models on 3 MRI-histology data subsets. First column: selection on 7 fixed mouse livers scanned *ex vivo* on a preclinical 9.4T Bruker system (**A** and **D**, left); Second column: selection on 6 liver tumours imaged *in vivo* on a clinical 1.5T Siemens system (**B** and **E**, middle); Third column: selection on 12 liver tumours imaged *in vivo* on a clinical 3T GE system (**C** and **F**, right). Plots on top (**A** to **C**) refer to dMRI model fitting performed on images where the vascular signal was suppressed (“whole image set fitting”, *b* > 1000 s/mm^2^ for suppression of PBS fluid within vessels on the 9.4T; *b* > 100 s/mm^2^ for IVIM signal suppression on clinical systems). Plots to the bottom (**D** to **F**) refer to dMRI model fitting performed on images where both vascular and extra-cellular, extra-vascular signals were suppressed (“high b-value fitting”, *b* > 1800 s/mm^2^ on the 9.4T; *b* > 900 s/mm^2^ on clinical systems). Violet: models where extra-cellular ADC is larger than intra-cellular ADC; orange: models with no constraints on which is larger between intra-/extra-cellular ADC. The *Bayesian Information Criterion* (BIC) selects a model depending on the goodness of model fitting. The *Histology Fidelity Criterion* (HFC) selects a model depending on the agreement between MRI volume-weighted Cell Size (*vCS*) and intra-cellular fraction (*F*) estimation with histology.

**Figure S4.**
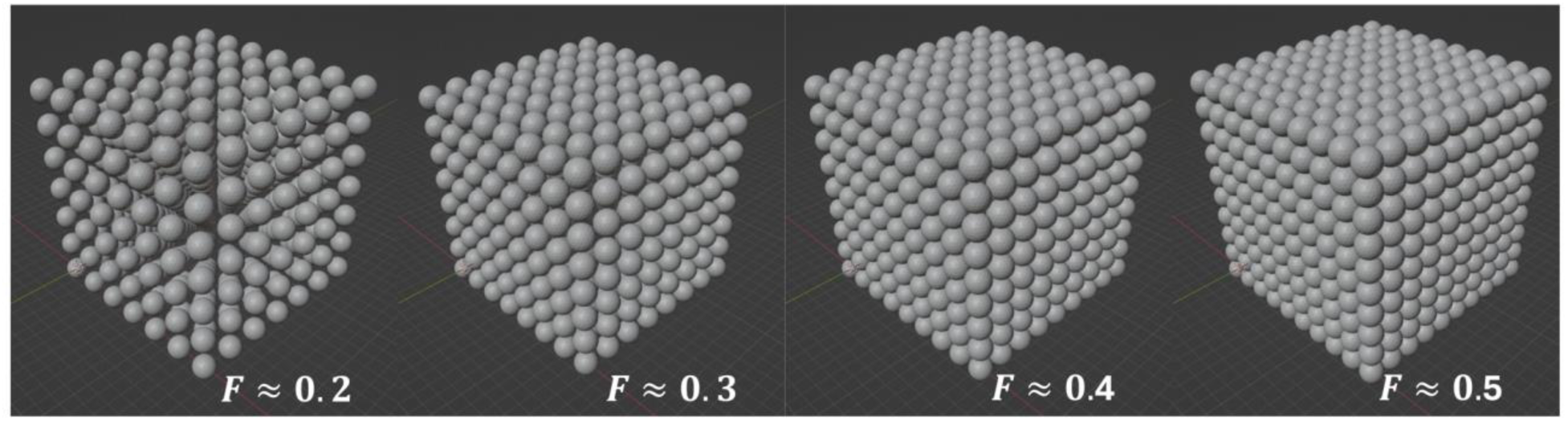
Environments used to generate synthetic dMRI signals in computer simulations. The synthetic environment consisted of meshed spheres of fixed diameter, representing cells, as this is a common biophysical model used in several dMRI techniques (e.g., VERDICT, IMPULSED). We used the synthetic environment to generate dMRI signals via Monte Carlo simulations for each of the 3 dMRI protocols considered in this study (the PGSE protocol used on the *ex vivo* mouse livers at 9.4T; the PGSE protocol used in patients *in vivo* at 3T; the DW TRSE protocol used in patients *in vivo* at 1.5T). Afterwards, we performed performed dMRI model selection on the synthetic signals, following the same procedures implemented for actual *ex vivo* and *in vivo* dMRI data. We controlled the intra-sphere fraction *F* by adding gaps of increasing size in-between abutting spheres packed in an ideal cubic lattice. We probed 4 different values of *F* (approximately equal to 0.197, 0.323, 0.406, 0.523; notice that the maximum theoretical value of *F* for cubic lattice packing is equal to 0.5236). For each value of *F*, we varied the cell diameter (8, 16, 22 and 30 µm), intra-sphere diffusivity (10 linearly-spaced values in the ranges [0.8; 2.6] µm^2^ ms^-1^ and [0.8; 3.0] µm^2^ ms^-1^ for the *ex vivo* and *in vivo* protocols respectively) and extra-sphere intrinsic diffusivity (again, 10 linearly-spaced values in the ranges [0.8; 2.6] µm^2^ ms^-1^ and [0.8; 3.0] µm^2^ ms^-1^ for the *ex vivo* and *in vivo* protocols respectively), generating a total of 1600 synthetic voxels. Before dMRI signal model fitting, we corrupted synthetic signal with Rician noise at a signal-to-noise ratio (SNR) of 30 on the b = 0 signal *s*(*b=0*) (SNR = *s*(*b=0*)/*σ*, where *σ* is the noise standard deviation).

**Figure S5.**
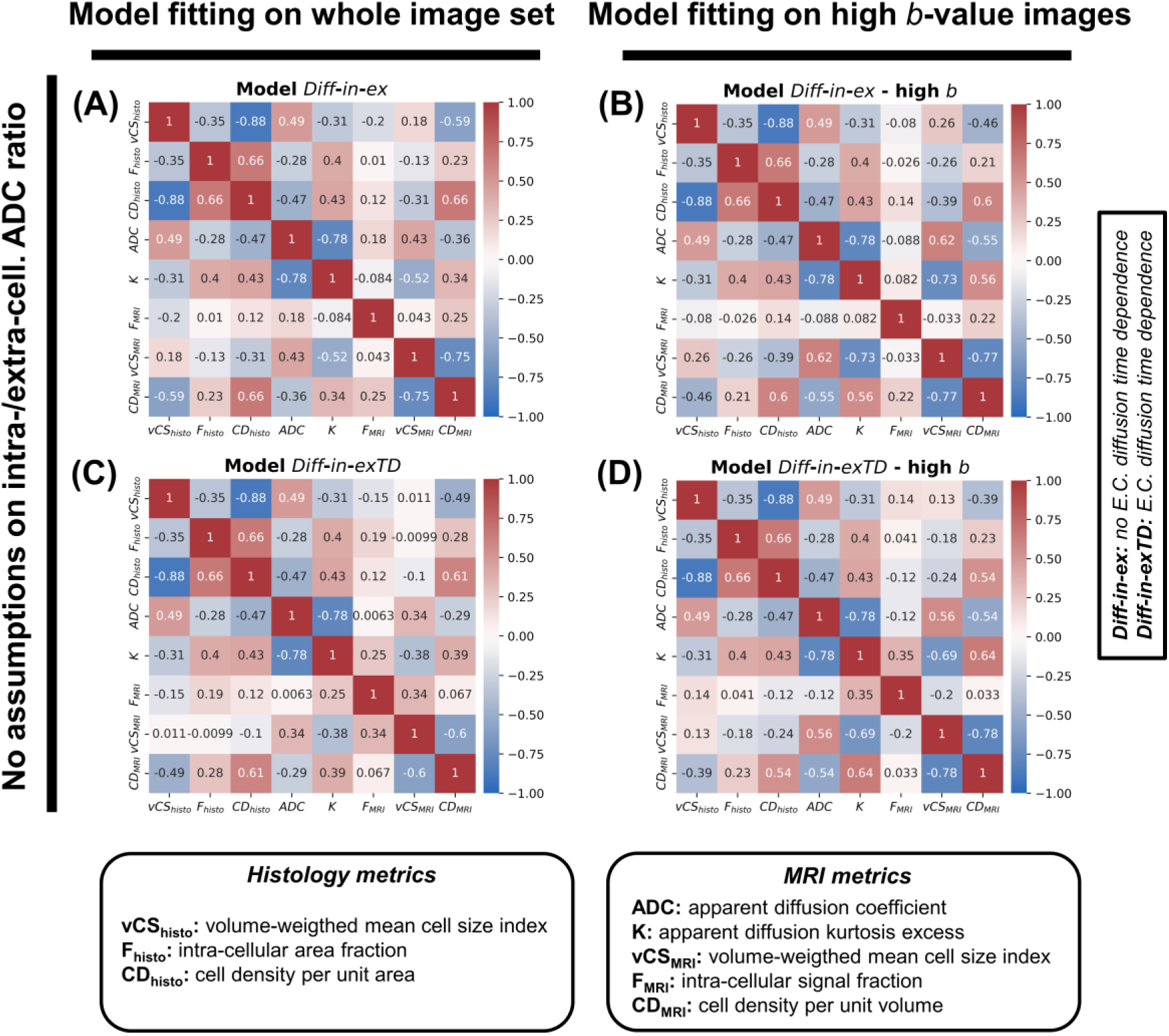
MRI-histology correlations for models with no assumptions on which is larger between intra-cellular and extra-cellular ADC. Matrices illustrating Pearson’s correlation coefficients among all possible pairs of MRI and histology metrics. Histological metrics are: intra- cellular area fraction *F*_*histo*_; volume-weighted mean cell size index *VCS*_*histo*_; cell density per unit area *CD*_*histo*_. MRI metrics are: apparent diffusion coefficient *ADC*; apparent diffusion excess kurtosis *K*; intra-cellular area fraction *F*_*MRI*_; volume-weighted mean cell size index *VCS*_*MRI*_; cell density per unit area *CD*_*MRI*_. Metrics *F*_*MRI*_, *VCS*_*MRI*_and *CD*_*MRI*_were obtained by fitting models with no assumptions on which is larger between intra-cellular and extra-cellular ADC (*Diff-in-ex* and *Diff-in-exTD*). The 4 panels refer to models *Diff-in-ex* and *Diff-in-exTD* fitted according to 2 different strategies. Panel (**A**): model *Diff-in-ex* fitted on the whole set of measurements with vascular signal suppression (*b* > 100 s/mm^2^ *in vivo*, *b* > 1000 s/mm^2^ *ex vivo*); panel (**B**): model *Diff-in-ex* fitted on high b-value measurements (*b* > 900 s/mm^2^ *in vivo*, *b* > 1800 s/mm^2^ *ex vivo*); panel (**C**): model *Diff-in-exTD* fitted on the whole set of measurements with vascular signal suppression; panel (**D**): model *Diff-in-exTD* fitted on high b-value measurements. We calculated correlation coefficients using a sample size of N = 25.

**Figure S6.**
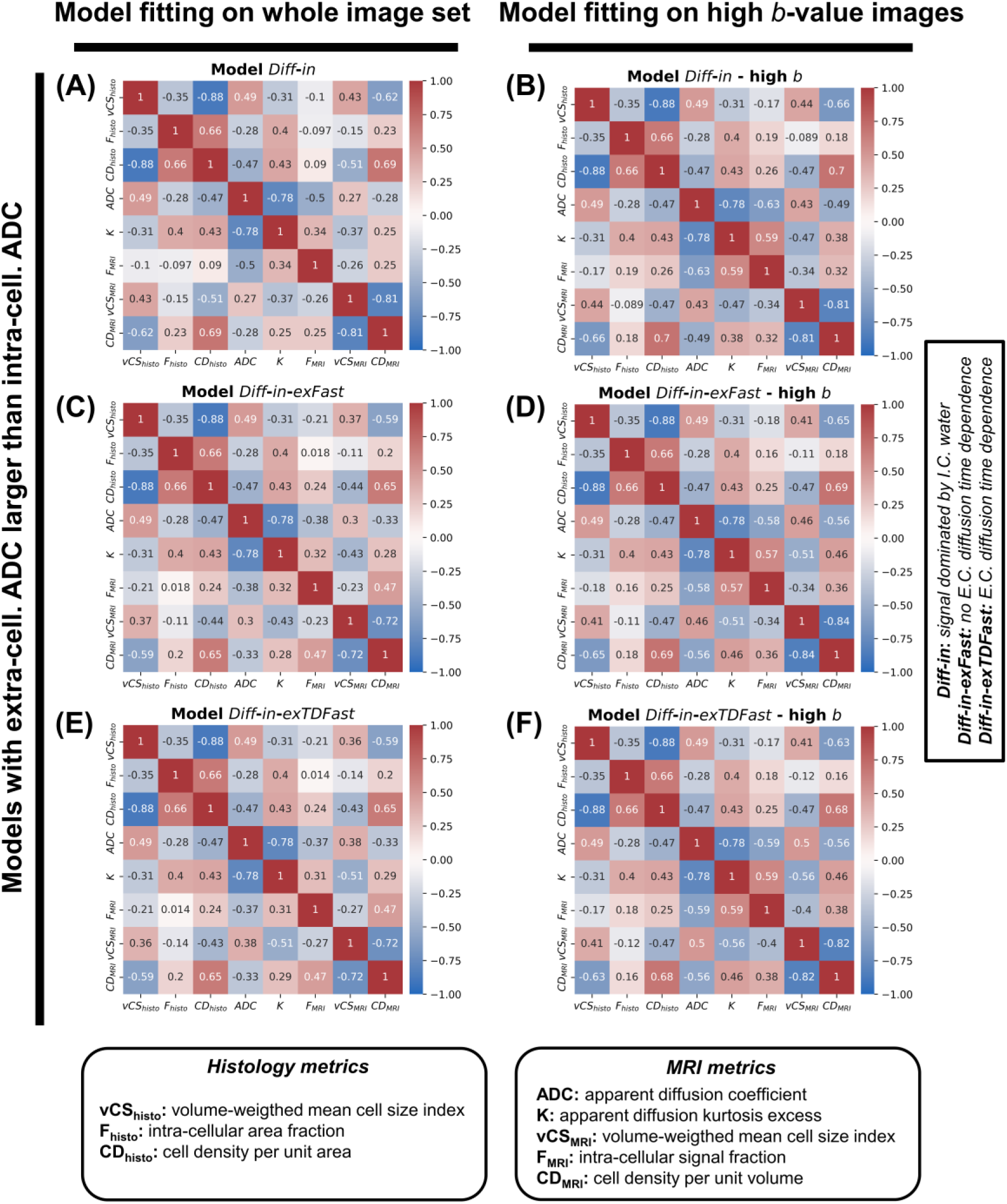
MRI-histology correlations for models where the extra-cellular ADC is constrained to be larger than the intra-cellular ADC. Matrices illustrating Pearson’s correlation coefficients among all possible pairs of MRI and histology metrics. Histological metrics are: intra-cellular area fraction *F*_*histo*_; volume-weighted mean cell size index *VCS*_*histo*_; cell density per unit area *CD*_*histo*_. MRI metrics are: apparent diffusion coefficient *ADC*; apparent diffusion excess kurtosis *K*; intra-cellular area fraction *F*_*MRI*_; volume-weighted mean cell size index *VCS*_*MRI*_; cell density per unit area *CD*_*MRI*_. Metrics *F*_*MRI*_, *VCS*_*MRI*_ and *CD*_*MRI*_ were obtained by fitting models that assume that the extra-cellular ADC is always larger than the intra-cellular ADC (*Diff-in*, *Diff-in-exFast* and *Diff-in-exTDFast*). The 6 panels refer to models *Diff-in*, *Diff-in-exFast* and *Diff-in-exTDFast* fitted according to 2 different strategies. Panel (**A**): model *Diff-in* fitted on the whole set of measurements with vascular signal suppression (*b* > 100 s/mm^2^ *in vivo*, *b* > 1000 s/mm^2^ *ex vivo*); panel (**B**): model *Diff-in* fitted on high b-value measurements (*b* > 900 s/mm^2^ *in vivo*, *b* > 1800 s/mm^2^ *ex vivo*); panel (**C**): model *Diff-in-exFast* fitted on the whole set of measurements with vascular signal suppression; panel (**D**): model *Diff-in-exFast* fitted on high b-value measurements; panel (**E**): model *Diff-in-exTDFast* fitted on the whole set of measurements with vascular signal suppression; panel (**F**): model *Diff-in-exTDFast* fitted on high b-value measurements. We calculated correlation coefficients using a sample size of N = 25.

**Figure S7.**
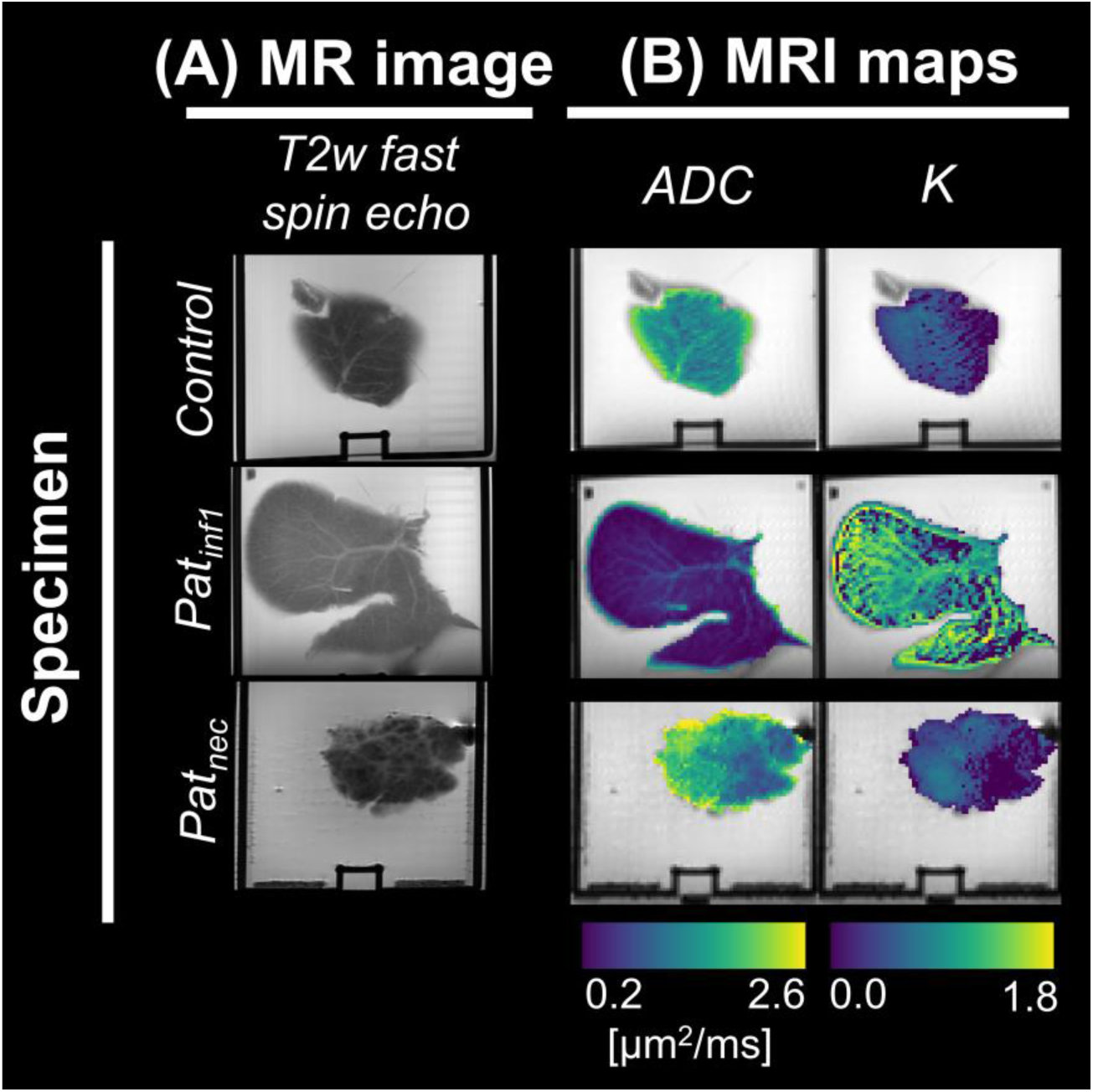
Standard DKI diffusion MRI metrics in fixed mouse livers *ex vivo*. **(A)**: high-resolution fast spin echo scan acquired in fixed mouse livers scanned *ex vivo* on the 9.4T Bruker system. **(B)**: standard diffusion metrics, namely *ADC* (apparent diffusion coefficient) and *K* (apparent diffusion kurtosis excess) from DKI. These metrics were obtained by fitting the standard diffusion kurtosis signal representation *s* = *s_0_* exp(*– b ADC + K (b ADC)^2^/6*) to the set of measurements at fixed TE = 45 ms and Δ = 30 ms. From top to bottom, the figure reports maps from 3 specimens, representative of the 3 different microstructural phenotypes seen in our mouse liver data. These are: normal liver structures (illustrated by the *Control* case, e.g., mouse with no biopsy implantation); pathology following biopsy implantation, consisting of an immature, lympho-proliferative process (infiltration of small cells in sinusoidal spaces, illustrated by case *Pat_inf_*); pathology following biopsy implantation, consisting of necrosis and inflammation (illustrated by case *Pat_nec_*).

**Figure S8.**
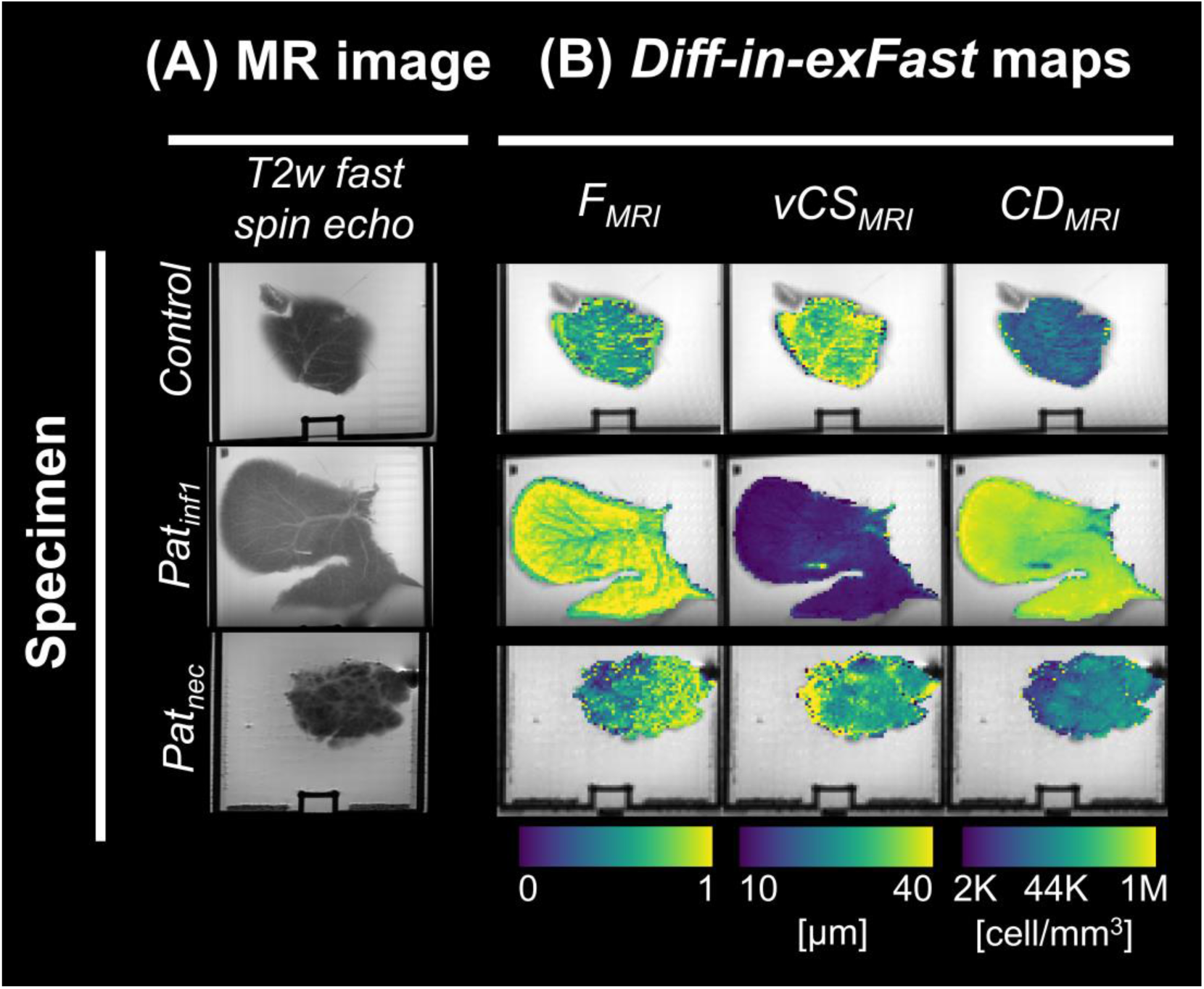
Key parametric maps of the *Diff-in-exFast* model on *ex vivo* mouse livers. **(A)**: high-resolution fast spin echo scan acquired in fixed mouse livers scanned *ex vivo* on the 9.4T Bruker system. **(B)**: metrics from the *Diff-in-exFast* model fitted to the whole DW image set (b-values with negligible vascular signal contributions, i.e., *b* > 1000 s/mm^2^ on fixed *ex vivo* tissue, to suppress signal from PBS-filled vessels). From left to right: intra-cellular signal fraction *F*_*MRI*_; volume-weighted cell size index *VCS*_*MRI*_index; cell density per unit volume *CD*_*MRI*_. Maps from 3 specimens are reported along different rows. The specimens are representative of the 3 different microstructural phenotypes seen in our mouse liver data. From top to bottom, these are: normal liver structures (illustrated by the *Control* case, e.g., mouse with no biopsy implantation); pathology following biopsy implantation, consisting of an immature, lympho-proliferative process (infiltration of small cells in sinusoidal spaces, illustrated by case *Pat_inf_*); pathology following biopsy implantation, consisting of extended necrosis and inflammation (illustrated by case *Pat_nec_*).

**Figure S9.**
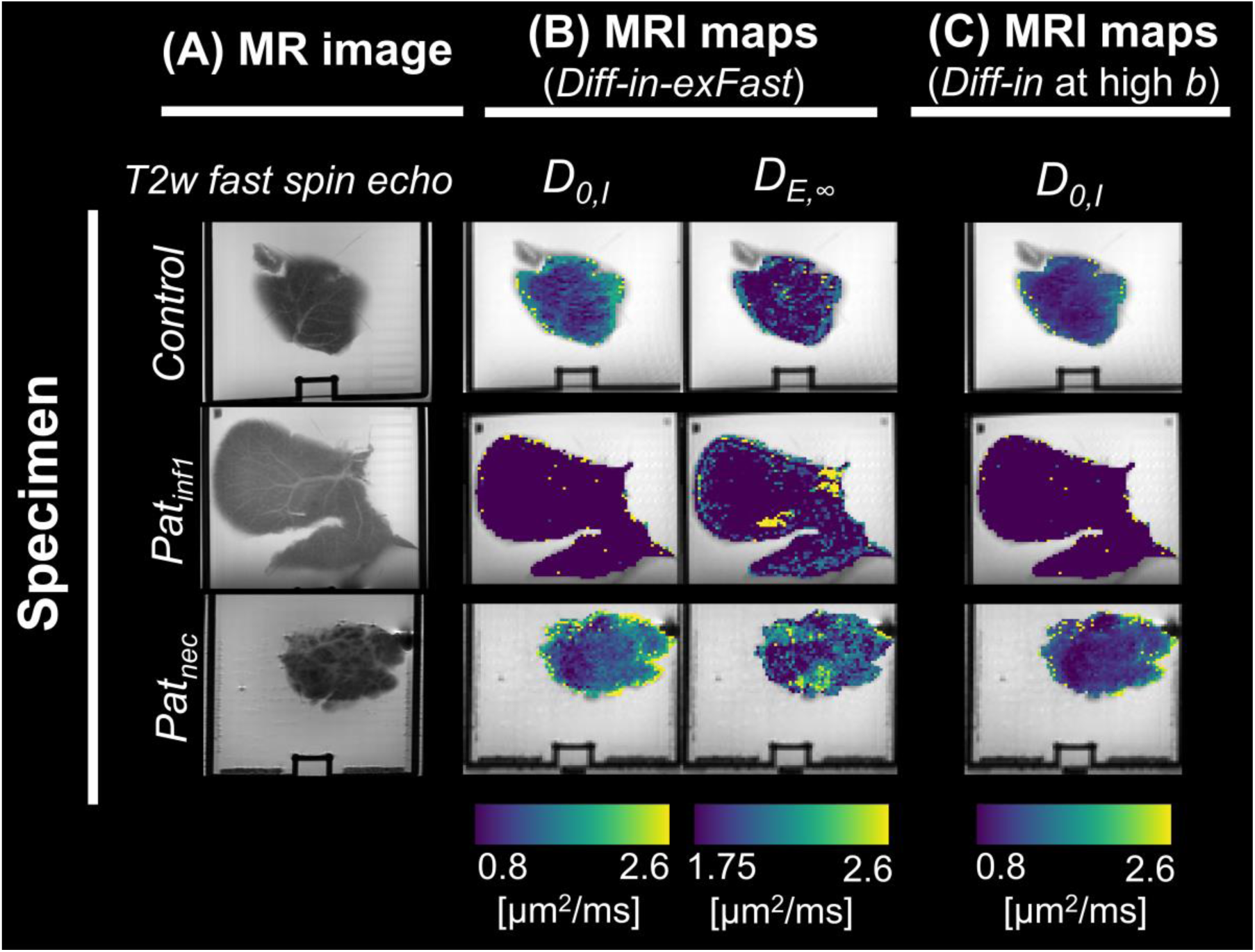
Diffusivity metrics from biophysical MRI models in fixed *ex vivo* mouse livers. **(A)**: high-resolution fast spin echo scan acquired in fixed mouse livers scanned *ex vivo* on the 9.4T Bruker system. **(B)**: diffusivity metrics from biophysical model *Diff-in-exFast*, namely: intrinsic intra-cellular cytosolic diffusivity *D*_0,*I*_; asymptotic extra-cellular diffusion coefficient *D*_*E*,∞_. **(C)**: intrinsic intra-cellular cytosolic diffusivity *D*_0,*I*_from model *Diff-in* fitted to high b-value images (*b* > 1800 s/mm^2^). Maps from 3 specimens are reported along different rows. The specimens are representative of the 3 different microstructural phenotypes seen in our mouse liver data. From top to bottom, these are: normal liver structures (illustrated by the *Control* case, e.g., mouse with no biopsy implantation); pathology following biopsy implantation, consisting of an immature, lympho-proliferative process (infiltration of small cells in sinusoidal spaces, illustrated by case *Pat_inf_*); pathology following biopsy implantation, consisting of extended necrosis and inflammation (illustrated by case *Pat_nec_*).

**Figure S10.**
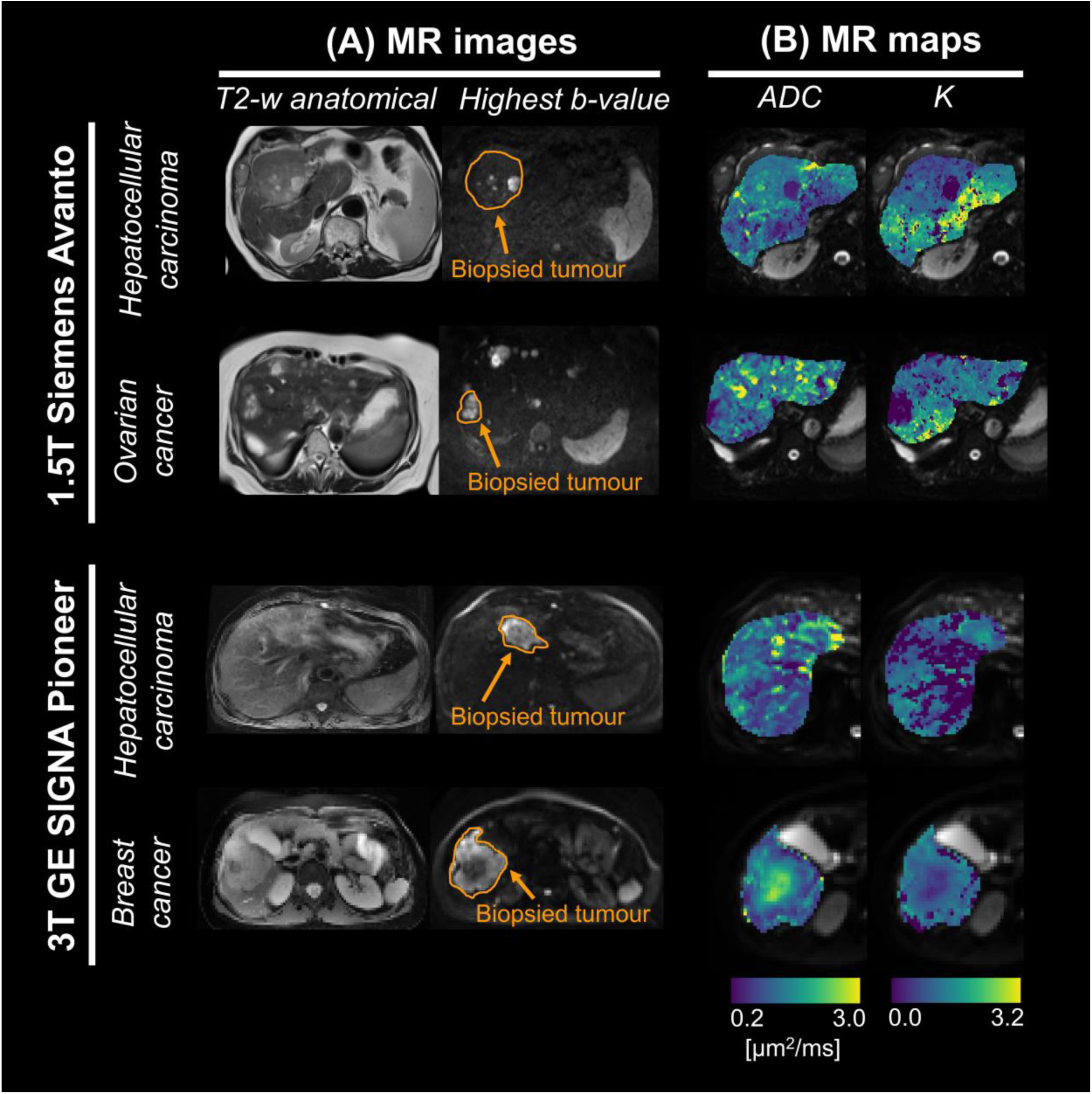
Standard DKI diffusion MRI metrics in patients *in vivo*. **(A)**: high-resolution fast spin echo scan as well as a high b-value diffusion image, with biopsied tumour outlined. **(B)**: standard DKI diffusion metrics in the biopsied tumour. Metrics are: apparent diffusion coefficient (*ADC*) and apparent diffusion kurtosis excess (*K*). These were obtained by fitting the standard diffusion kurtosis signal representation *s* = *s_0_* exp(*– b ADC + K (b ADC)^2^/6*) to the set of measurements at fixed, minimum TE and *b* > 100 s/mm^2^. Maps are shown in four representative patients (two patients for each MRI scanner), along different rows. For the 1.5T Siemens scanner (first and second rows from top): a primary hepatocellular carcinoma case and a patient with liver metastases from ovarian cancer. For the 3T GE scanner (third and fourth rows from top): a primary hepatocellular carcinoma case and a patient with liver metastases from breast cancer.

**Figure S11.**
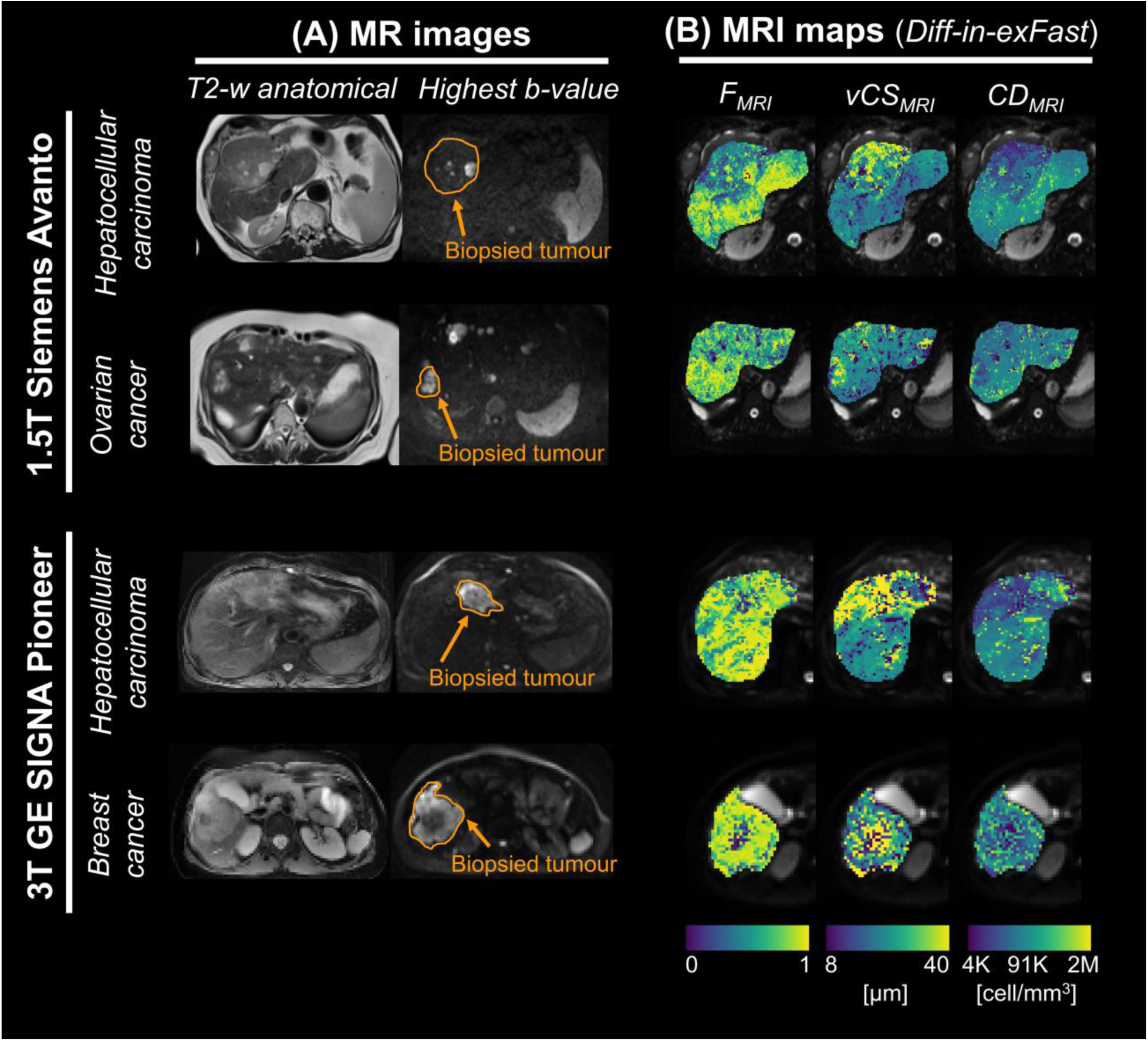
Key parametric maps of the *Diff-in-exFast* model in patients *in vivo*. **(A)**: high-resolution fast spin echo scan as well as a high b-value diffusion image, with biopsied tumour outlined. **(B)**: salient metrics of the *Diff-in-exFast* model fitted to the whole set of images with negligible vascular signal contributions (*b* > 100 s/mm^2^). Metrics are shown in the biopsied tumour. From left to right: intra-cellular signal fraction *F*_*MRI*_; volume-weighted cell size index *VCS*_*MRI*_ index; cell density per unit volume *CD*_*MRI*_. Metrics are shown in four representative patients (two patients for each MRI scanner), along different rows. For the 1.5T Siemens scanner (first and second rows from top): a primary hepatocellular carcinoma case and a patient with liver metastases from ovarian cancer. For the 3T GE scanner (third and fourth rows from top): a primary hepatocellular carcinoma case and a patient with liver metastases from breast cancer.

**Figure S12.**
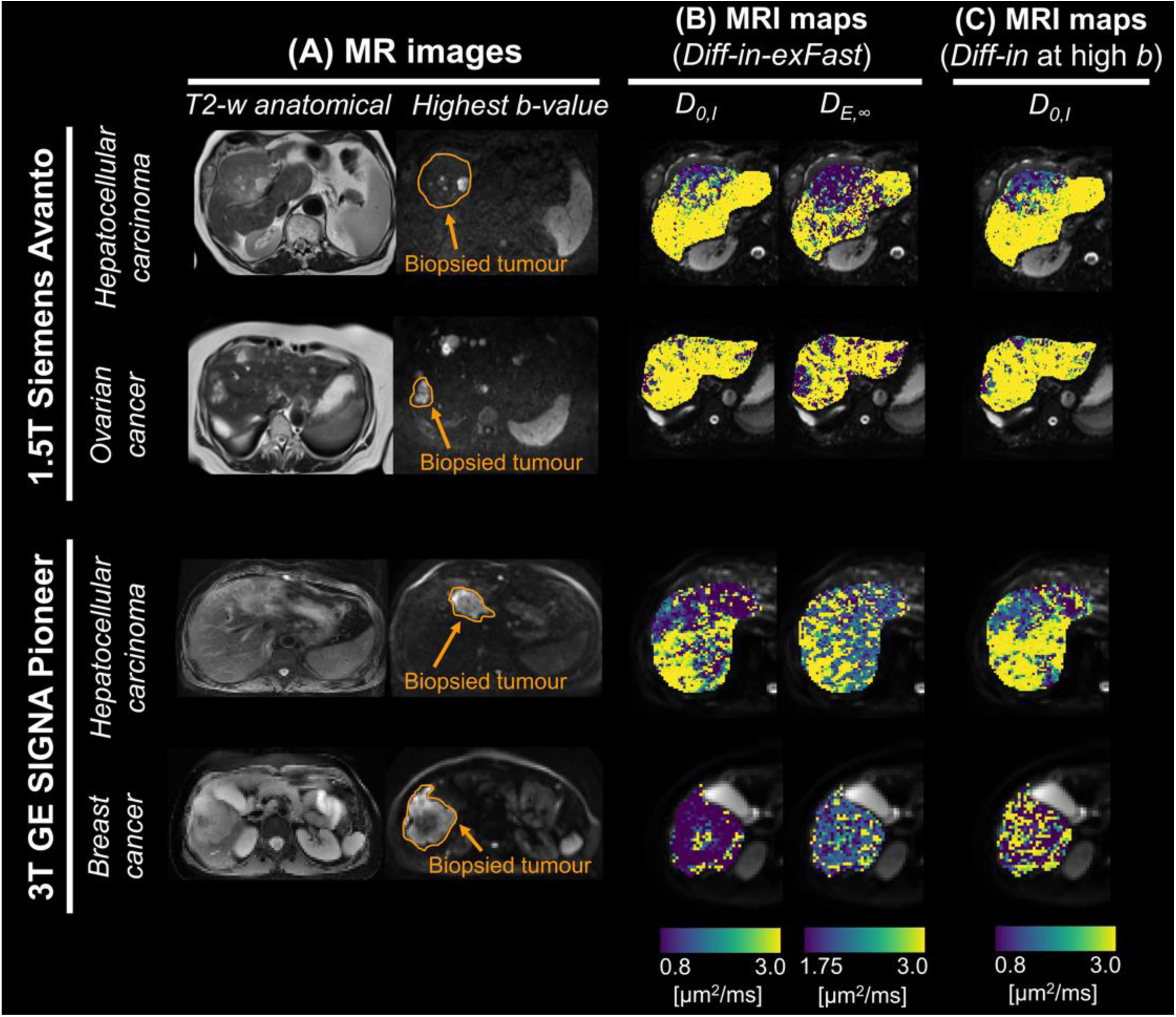
Diffusivity metrics from biophysical MRI models in patients *in vivo*. **(A)**: high-resolution fast spin echo scan as well as a high b-value diffusion image, with biopsied tumour outlined. **(B)**: diffusivity maps from biophysical model *Diff-in-exFast* in the biopsied tumour. Metrics are: intra-cellular cytosolic diffusivity *D*_0,*I*_ and asymptotic extra-cellular diffusion coefficient *D*_*E*,∞_. **(C)**: intra-cellular cytosolic diffusivity *D*_0,*I*_for biophysical model *Diff-in* fitted being fitted only to high b-value images (*b* > 900 s/mm^2^). Metrics are shown in four representative patients (two patients for each MRI scanner), along different rows. For the 1.5T Siemens scanner (first and second rows from top): a primary hepatocellular carcinoma case and a patient with liver metastases from ovarian cancer. For the 3T GE scanner (third and fourth rows from top): a primary hepatocellular carcinoma case and a patient with liver metastases from breast cancer.

**Figure S13.**
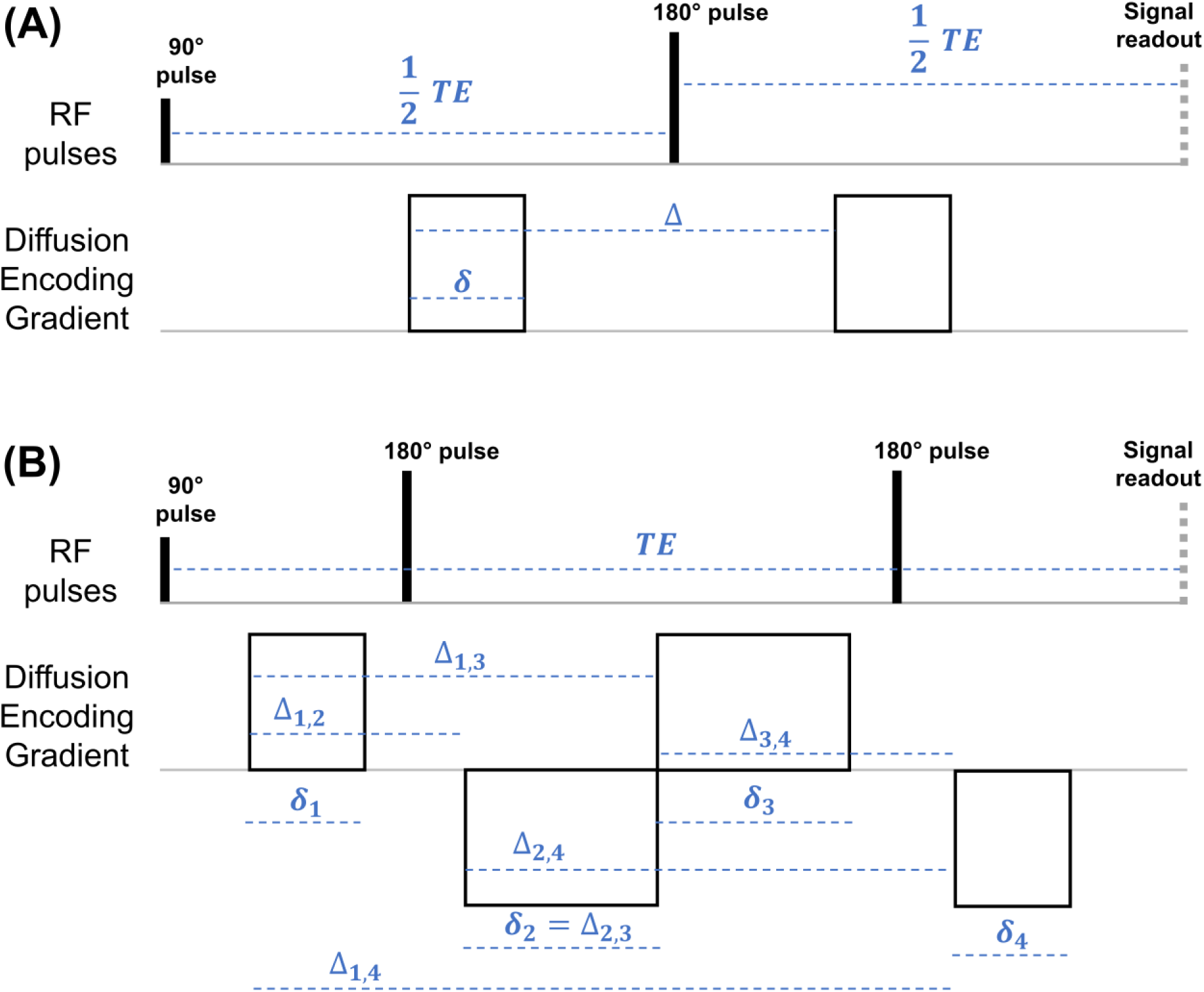
Schematic of the dMRI sequences used in this study. (**A**): pulsed gradient spin echo (PGSE sequence, also known as Stejskal-Tanner sequence, pulsed-field gradient (PFG), or single linear diffusion encoding) used to acquire data on the 9.4T Bruker system on fixed mouse livers *ex vivo* and on the 3T GE system on patients *in vivo*. δ and Δ respectively indicate the diffusion gradient duration and separation, while TE is the echo time. (**B**): twice-refocussed diffusion-weighted spin echo sequence used to acquire data on the 1.5T Siemens system on patients *in vivo*. δ*_n_* and Δ*_n,m_* respectively indicate the duration of the *n*-th gradient lobe and the separation time between the *n*-th and *m*-th gradient lobes, for *n,m* = 1, …, 4. TE is again the echo time. In both panels, “Signal readout” corresponds to sampling the center of the k-space (i.e., zero spatial frequency).

**Table S1:** results of the model selection based on the Total Correlation Score (TCS) as obtained on simulated dMRI signals. We performed model selection on synthetic signals simulated for all the dMRI protocols considered in this study (*ex vivo* PGSE, used on fixed mouse livers; *in vivo* PGSE and DW TRSE, used in patients *in vivo*; see Methods for a full description of the protocols). We fitted the models on protocol subsets obtained with the same b-value thresholds used when analysing actual MRI signals (“Regular fit”: fitting on all b-values with negligible vascular contributions; “High *b* only fit”: fitting on b-values minimising extra-cellular signal contributions). The Table reports the value of *TCS* = *r*(*vCS_est_,vCS_gt_*) + *r*(*F_est_,F_gt_*), where *vCS* is the cell size, *F* the intra-cellular fraction, *r*(*x,y*) the Pearson’s correlation between variables *x* and *y* computed pooling together all synthetic voxels, and where subscripts *est* and *gt* respectively indicate estimated and ground truth values. Higher values of *TCS* point towards better model performance. For each protocol and fitting strategy, the model with the highest *TCS* is flagged by gray shadowing and bold font.

|  | Protocol:<br>*ex vivo* |  | Protocol:<br>*in vivo* PGSE |  | Protocol:<br>*in vivo* TRSE |  |
| --- | --- | --- | --- | --- | --- | --- |
|  | Regular<br>fit | High *b*<br>only fit | Regular<br>fit | High *b*<br>only fit | Regular<br>fit | High *b*<br>only fit |
| Model |  |  |  |  |  |  |
| *Diff-in-exTD* | 0.217 | −0.111 | 0.312 | 0.202 | 0.604 | 0.483 |
| *Diff-in-ex* | 0.406 | 0.089 | 0.472 | 0.335 | **0.700** | 0.550 |
| *Diff-in-exTDFast* | 0.948 | 0.827 | 0.536 | 0.336 | 0.618 | 0.544 |
| *Diff-in-exFast* | 0.952 | 0.850 | 0.563 | 0.349 | 0.626 | 0.547 |
| *Diff-in* | **1.222** | **0.977** | **0.773** | **0.462** | 0.543 | **0.630** |

**Table S2.** Results of the model selection based on the Histology Fidelity Criterion (HFC) as obtained on simulated dMRI signals. We performed model selection on synthetic signals simulated for all the dMRI protocols considered in this study (*ex vivo* PGSE, used on fixed mouse livers; *in vivo* PGSE and DW TRSE, used in patients *in vivo*; see Methods for a full description of the protocols). We fitted the models on protocol subsets obtained with the same b-value thresholds used when analysing actual MRI signals (“Regular fit”: fitting on all b-values with negligible vascular contributions; “High *b* only fit”: fitting on b-values minimising extra-cellular signal contributions). For each model, the table reports the percentage of synthetic voxels where *HFC* = |*vCS_est_ – vCS_gt_*|/*vCS_gt_* + |*F_est_ – F_gt_*|/*F_gt_* was the lowest across all models. Above, *vCS* is the cell size, *F* the intra-cellular fraction, and subscripts *est* and *gt* respectively indicate estimated and ground truth values. Higher percentages indicate smaller estimation errors, and therefore point towards better model performance. For each protocol and fitting strategy, the model with the highest proportion of synthetic voxels with minimum HFC is flagged by gray shadowing and bold font.

|  | Protocol:<br>*ex vivo* |  | Protocol:<br>*in vivo* PGSE |  | Protocol:<br>*in vivo* TRSE |  |
| --- | --- | --- | --- | --- | --- | --- |
|  | Regular<br>fit | High *b*<br>only fit | Regular<br>fit | High *b*<br>only fit | Regular<br>fit | High *b*<br>only fit |
| Model |  |  |  |  |  |  |
| *Diff-in-exTD* | 12.62% | 13.56% | 21.69% | 22.10% | 26.69% | 20.69% |
| *Diff-in-ex* | 21.19% | 16.62% | **25.94%** | 20.35% | **28.94%** | 24.31% |
| *Diff-in-exTDFast* | 15.50% | 14.94% | 10.75% | 12.27% | 19.69% | 14.69% |
| *Diff-in-exFast* | 21.81% | 10.50% | 22.25% | 11.33% | 19.12% | 12.12% |
| *Diff-in* | **28.88%** | **44.38%** | 19.38% | **33.94%** | 5.56% | **28.19%** |

**Table S3.** Results of the model selection based on the Bayesian Information Criterion (BIC) as obtained on simulated dMRI signals. We performed model selection on synthetic signals simulated for all the dMRI protocols considered in this study (*ex vivo* PGSE, used on fixed mouse livers; *in vivo* PGSE and DW TRSE, used in patients *in vivo*; see Methods for a full description of the protocols). We fitted the models on protocol subsets obtained with the same b-value thresholds used when analysing actual MRI signals (“Regular fit”: fitting on all b-values with negligible vascular contributions; “High *b* only fit”: fitting on b-values minimising extra-cellular signal contributions). For each model, the table reports the percentage of synthetic voxels where the Bayesian Information Criterion (BIC, a standard metric of model fitting quality that penalises model complexity) was the lowest across all models. Higher percentages indicate smaller BIC values across synthetic voxels, and therefore point towards better model fitting quality. For each protocol and fitting strategy, the model with the highest proportion of synthetic voxels with minimum HFC is flagged by gray shadowing and bold font.

|  | Protocol:<br>*ex vivo* |  | Protocol:<br>*in vivo* PGSE |  | Protocol:<br>*in vivo* TRSE |  |
| --- | --- | --- | --- | --- | --- | --- |
|  | Regular<br>fit | High *b*<br>only fit | Regular<br>fit | High *b*<br>only fit | Regular<br>fit | High *b*<br>only fit |
| Model |  |  |  |  |  |  |
| *Diff-in-exTD* | 0.12% | 0.00% | 0.00% | 0.19% | 0.00% | 0.00% |
| *Diff-in-ex* | 31.25% | 15.88% | 17.44% | 4.69% | **40.69%** | 13.69% |
| *Diff-in-exTDFast* | 0.00% | 0.00% | 0.00% | 0.00% | 0.00% | 0.00% |
| *Diff-in-exFast* | 3.44% | 0.06% | 6.69% | 0.00% | 21.38% | 1.06% |
| *Diff-in* | **65.19%** | **84.06%** | **75.88%** | **95.12%** | 38.00% | **85.25%** |

**Table S4.** Descriptive statistics of histology and MRI metrics in the fixed mouse livers. The table reports mean and standard deviation (within brackets) of histology and dMRI metrics in the 7 fixed mouse livers that were scanned on a 9.4T Bruker system. Histological maps were computed within patches matching the in-plane MRI resolution and then warped non-linearly to dMRI space. The histological maps are: per-patch intra-cellular area fraction *F*_*histo*_, per-patch arithmetic mean cell size *aCS*_*histo*_; per-patch volume-weighted mean cell size *VCS*_*histo*_, cell density per unit patch area *CD*_*histo*_. dMRI metrics are: apparent diffusion coefficient *ADC*, apparent diffusion kurtosis excess *K*, intra-cellular signal fraction *F*_*MRI*_, volume-weighted cell size index *VCS*_*MRI*_, apparent cell density per unit volume *CD*_*MRI*_. Metrics *F*_*MRI*_, *VCS*_*MRI*_and *CD*_*MRI*_are reported for both models *Diff-in-exFast* and model *Diff-in*, with *Diff-in* fitted only to high b-value images (*b* > 1800 s/mm^2^). In model *Diff-in-exFast*, the extra-cellular ADC does not feature diffusion time dependence and is constrained to be larger than the intra-cellular ADC. In model *Diff-in*, the extra-cellular signal is modelled as negligible compared to the intra-cellular one (i.e., total signal dominated by intra-cellular water). Specimens are: *Control* (normal liver structures); *NA1* and *NA2* (normal appearing cases, i.e., normal liver structures despite sub-cutaneous biopsy implantation); *Pat_inf1-3_* (cases developing liver pathology following sub-cutaneous biopsy implantation, consisting of small cell infiltration in sinusoidal spaces, in between larger hepatocytes); *Pat_nec_* (case developing liver pathology following sub-cutaneous biopsy implantation, consisting of necrosis and inflammation). *aCS*_*histo*_, always considerably lower than *VCS*_*histo*_, was included to highlight the impact of the largest cells in the computation of statistics based on weighting by cell volume (*VCS*_*histo*_).

| Specimen | Histology | | | | Standard diffusion metrics | | *Diff-in-exFast* model | | | *Diff-in* model at high $b$ | | |
| --- | --- | --- | --- | --- | --- | --- | --- | --- | --- | --- | --- | --- |
| | $F_{histo}$ | $aCS_{histo}$<br>[$\mu m$] | $vCS_{histo}$<br>[$\mu m$] | $CD_{histo}/10^2$<br>[cell/mm<sup>2</sup>] | $ADC$<br>[$\mu m^2/ms$] | $K$ | $F_{MRI}$ | $vCS_{MRI}$<br>[$\mu m$] | $CD_{MRI}/10^5$<br>[cell/mm<sup>3</sup>] | $F_{MRI}$ | $vCS_{MRI}$<br>[$\mu m$] | $CD_{MRI}/10^5$<br>[cell/mm<sup>3</sup>] |
| *Control* | 0.75<br>(0.18) | 21.63<br>(1.85) | 27.1<br>(1.6) | 3.2<br>(1.1) | 1.50<br>(0.32) | 0.33<br>(0.20) | 0.60<br>(0.18) | 33.2<br>(5.7) | 0.38<br>(1.69) | 0.56<br>(0.11) | 36.5<br>(5.1) | 0.28<br>(1.44) |
| *NA1* | 0.59<br>(0.25) | 23.97<br>(3.05) | 27.8<br>(3.2) | 2.1<br>(1.1) | 1.61<br>(0.37) | 0.10<br>(0.15) | 0.66<br>(0.25) | 30.5<br>(8.4) | 0.69<br>(2.18) | 0.54<br>(0.16) | 32.2<br>(7.5) | 0.45<br>(1.68) |
| *NA2* | 0.76<br>(0.14) | 23.46<br>(1.41) | 29.4<br>(1.2) | 2.7<br>(0.7) | 1.43<br>(0.44) | 0.51<br>(0.29) | 0.80<br>(0.16) | 22.2<br>(4.8) | 1.01<br>(1.35) | 0.71<br>(0.12) | 22.8<br>(5.0) | 0.83<br>(1.15) |
| *Pat<sub>infl1</sub>* | 0.80<br>(0.20) | 15.73<br>(2.18) | 20.8<br>(2.7) | 6.7<br>(2.2) | 0.58<br>(0.41) | 0.98<br>(0.49) | 0.83<br>(0.13) | 13.4<br>(3.1) | 4.20<br>(2.22) | 0.77<br>(0.14) | 12.4<br>(3.0) | 4.86<br>(2.27) |
| *Pat<sub>infl2</sub>* | 0.79<br>(0.20) | 21.41<br>(2.29) | 26.8<br>(2.7) | 3.5<br>(1.2) | 1.67<br>(0.39) | 0.17<br>(0.17) | 0.37<br>(0.22) | 31.2<br>(7.5) | 0.44<br>(2.06) | 0.41<br>(0.14) | 37.1<br>(6.0) | 0.30<br>(1.57) |
| *Pat<sub>infl3</sub>* | 0.70<br>(0.27) | 20.95<br>(1.75) | 27.6<br>(1.7) | 3.1<br>(1.3) | 1.57<br>(0.62) | 0.43<br>(0.31) | 0.63<br>(0.23) | 23.6<br>(8.8) | 1.27<br>(2.62) | 0.59<br>(0.20) | 24.2<br>(8.0) | 0.97<br>(2.01) |
| *Pat<sub>nec</sub>* | 0.52<br>(0.25) | 19.10<br>(3.33) | 25.9<br>(2.8) | 3.4<br>(2.3) | 1.49<br>(0.44) | 0.31<br>(0.22) | 0.61<br>(0.23) | 28.2<br>(6.6) | 0.58<br>(1.93) | 0.54<br>(0.16) | 31.6<br>(6.7) | 0.42<br>(1.66) |

**Table S5.** Correlation between histological metrics and tumour volume. The table reports the Pearson’s correlation coefficient between histological metrics *F_histo_* (intr-cellular fraction), *vCS_histo_* (volume-weighted characteristic cell size), *aCS_histo_* (arithmetic mean cell size index), *CD_histo_* (cell density per unit area) and *F_Ki67_* (fraction of Ki67-labelled material) and the log_10_ of the volume of the tumour from which the biopsy was obtained. For this calculation, we did not include the two cases where the biopsy was obtained after starting immunotherapy (N = 16 for HE-derived metrics *F_histo_*, *vCS_histo_*, *aCS_histo_* and *CD_histo_*; N = 10 for *F_Ki67_*).

|  | **Correlation with tumour volume** |
| --- | --- |
| $F_{histo}$ | $r = -0.102, p = 0.708$ |
| $vCS_{histo}$ | $r = -0.038, p = 0.890$ |
| $aCS_{histo}$ | $r = -0.077, p = 0.776$ |
| $CD_{histo}$ | $r = 0.001, p = 0.997$ |
| $F_{Ki67}$ | $r = -0.306, p = 0.389$ |

